# Innate immune cytokine profiling and biomarker identification for outcome in dengue patients

**DOI:** 10.1101/2020.10.14.20212001

**Authors:** Sai Pallavi Pradeep, Pooja Hoovina Venkatesh, Nageswar R. Manchala, Arjun Vayal Veedu, Rajani K. Basavaraju, Leela Selvasundari, Manikanta Ramakrishna, Yogitha Chandrakiran, Vishwanath Krishnamurthy, Shivaranjani Holigi, Tinku Thomas, Cecil R. Ross, Mary Dias, Vijaya Satchidanandam

## Abstract

Biomarkers of progression to severe dengue are urgently required for effective patient management. Innate immune cells have been implicated in the enhancement of infection and “cytokine storm” associated with dengue severity. Using intracellular cytokine staining and flow cytometry, we observed significantly higher proportions of innate immune cells secreting inflammatory cytokines dominated by IFN-γ and TNF-α at admission associated with good prognosis. Secondary dengue predisposed to severe outcomes. In patients with severe dengue and those with liver impairment, early activation as well as efficient down-regulation of innate responses were compromised. IFN-γ^+^CD56^+^CD3^+^ NKT cells and IL-6^+^ granulocytes served as novel biomarkers of progression to severity (composite AUC=0.85-0.9). Strong correlations among multiple cytokine-secreting innate cell subsets pointed to coordinated activation of the entire innate immune system by DENV.

**One Sentence Summary:** Activation and efficient attenuation of innate immunity are both compromised in severe dengue.

## Main Text

Dengue virus (DENV) afflicts ∼130 million people annually with 70% contribution from Asia (*1*). Majority of patients present with mild dengue fever (DF) while ∼5-20% develop severe dengue (SD)-characterized by plasma leakage leading to shock and/or organ impairment that predominantly manifest during defervescence (*2, 3*). The need to identify severe cases early, to facilitate interventions that reduce mortality has spawned numerous investigations to find biomarkers of severity. The phenomenon of antibody dependent enhancement (ADE) of viral infectivity has been implicated in dengue pathology (*4, 5*) with monocytes that both support virus growth and express all three classes of Fc-receptors (FcγR, FcεR and FcαR) being major contributors (*6, 7*). Deficits in T cell signaling and cytokine secretion have been reported in dengue patients from Sri Lanka, India and Thailand (*8, 9*). Combined with spontaneous high production of multiple cytokines from unstimulated PBMCs (*9*), this implicated innate cells as likely sources of the ‘cytokine storm’ in SD (*10*), supported by in vitro assays of infected monocytes and natural killer (NK) cells (*11, 12*). Conflicting patterns for serum levels of TNF-α, IP-10 and IFN-γ as a function of severity were reported (*13-16*), perhaps due to variation in sampling time following infection. The failure to exploit flow cytometry made it impossible to identify the source of cytokines measured by ELISA. Against this backdrop, we carried out a blinded study (fig. S1) to query the secretion of cytokines by human innate immune cells in response to DENV using intracellular cytokine staining and flow cytometry (fig. S2 & S3).

Dengue infection, in contrast to fevers from other etiologies, caused a surge in secretion of inflammatory cytokines from a variety of innate cells (Fig. 1, A-F). TNF-α was by far the most abundant cytokine secreted by multiple cell subsets from a vast majority of patients while an impressive 95.5% of patients secreted IFN-γ from CD56^+^CD3^+^ NKT cells (table S1). We also observed significantly greater numbers of dual-functional IL-10^+^TNF-α^+^ granulocytes and IFN-γ^+^TNF-α^+^CD56^+^CD3^+^ NKT cells, in dengue patients than in febrile controls (fig. S4A, B). Polyfunctional CD56^+^CD3^+^ NKT cells among others, could indeed be visualized by t-Distributed Stochastic Neighbor Embedding analysis (fig. S5). Thus, in addition to monocytes which support dengue replication (*11*), all other innate cell subsets were also activated by DENV. As reported previously (*17*), DENV infection significantly increased the numbers of CD14^+^ and CD14^+^CD16^+^ monocytes and significantly reduced CD56^+^CD3^+^ NKT cell numbers compared to controls (table S2; fig. S6A, B). Some cell subsets progressively expanded with increase in disease severity (table S3). CD14^+^CD16^+^ intermediate monocytes or CD19^+^ B cells were the highest per cell secretors of all cytokines (fig. S7, A-E; table S1). The median florescence intensity (MFI) was comparable across severity except for TNF-α^+^CD56^+^CD3^+^ NKT cells (fig. S7F, G).

**Fig. 1.**
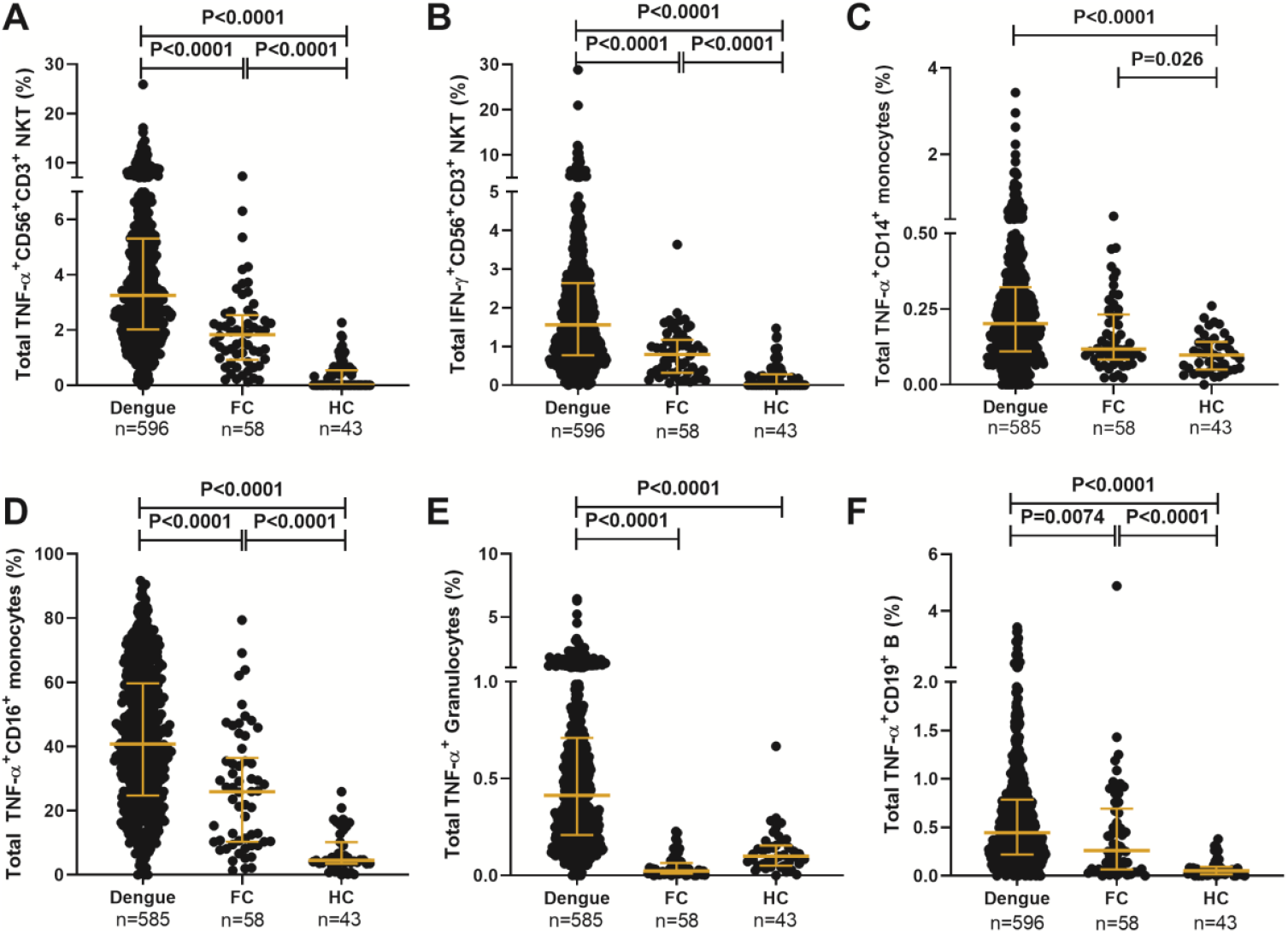
DENV activates cytokine secretion from innate immune cells. Frequency of total (**A**) TNF-α^+^CD56^+^CD3^+^- and (**B**) IFN-γ^+^CD56^+^CD3^+^-NKT cells, (**C**) TNF-α^+^CD14^+^ monocytes, (**D**) TNF-α^+^CD16^+^ monocytes, (**E**) TNF-α^+^ granulocytes and (**F**) TNF-α^+^CD19^+^ B cells from dengue patients compared to febrile controls (FC) and healthy controls (HC). Each dot represents one patient; P value displayed with median and IQR.

When assessed as a function of disease severity, a significantly greater proportion of TNF-α^+^ granulocytes were evident in DF relative to DFWS/SD (Fig. 2A), a trend also evident for dual secretion of TNF-α and IL-6 by this cell subset and IL-10 secretion by CD56^+^CD3^+^ NKT cells (Fig. 2B, C). When we used bleed-scores or liver enzyme levels as a surrogate of severity, those with no bleeding or normal liver enzyme levels carried a significantly greater proportion of innate cells secreting inflammatory cytokines compared to those with varying degrees of hemorrhage or abnormal liver enzyme levels (Fig. 2, D-F). As expected, the odds of severity were greater in patients with high liver enzymes (fig. S8A, B) and secondary dengue (fig. S8C, D). The latter predisposition was accompanied by a significantly lower proportion of TNF-α secreting monocytes, granulocytes and CD19^+^ B cells (fig. S8, E-H) reinforcing the reported (*4, 5*) deleterious role of pre-existing immunity on dengue outcomes. In addition, age and gender also influenced cytokine secretion from monocyte subsets (fig. S8I, J). Despite variations within the cohort based on gender, age and primary/secondary dengue, the higher percentages of innate cell subsets secreting inflammatory cytokines, impressively correlated with better prognosis as suggested by earlier blood transcriptome studies (*18*). In contrast, higher frequency of monofunctional IP-10^+^CD19^+^ B cells was associated with severity (fig. S9A, B).

**Fig. 2.**
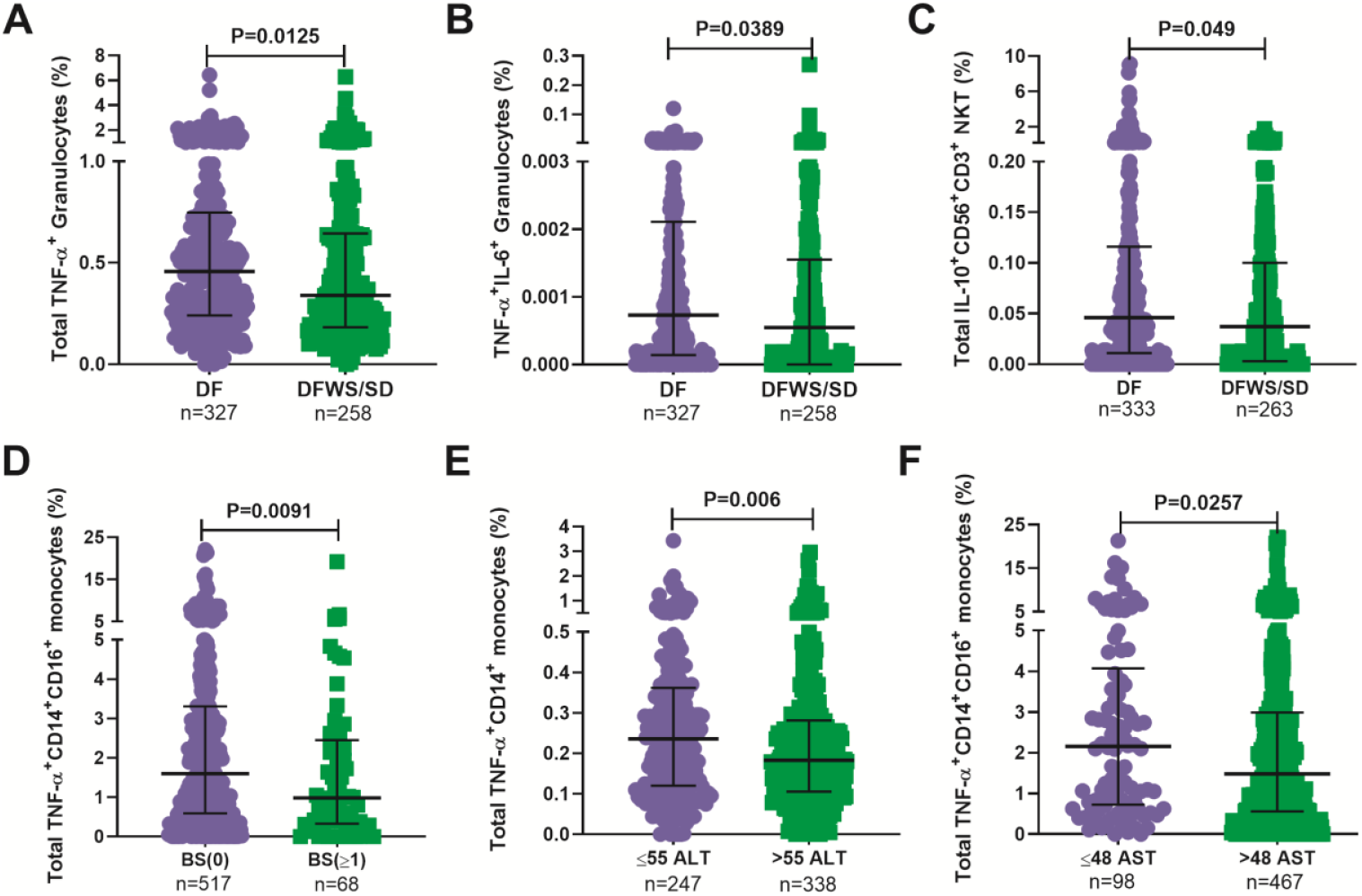
Higher percentages of innate immune cells secreting inflammatory cytokines are associated with better prognosis. Frequency of (**A**) total TNF-α^+^ granulocytes, (**B**) TNF-α^+^IL-6^+^ granulocytes, and (**C**) total IL-10^+^CD56^+^CD3^+^ NKT cells compared between DF and DFWS/SD. (**D**) Frequency of total TNF-α^+^CD14^+^CD16^+^ monocytes compared between bleed-scores (BS). Frequency of total (**E**) TNF-α^+^CD14^+^ monocytes and (**F**) TNF-α^+^CD14^+^CD16^+^ monocytes compared between normal and elevated ALT (>55 IU/L; E) and AST (>48 IU/L; F). P values with median and IQR reported.

Total TNF-α^+^ percentages within all subsets displayed strong positive correlation with one another (r=0.63-0.77), while IFN-γ^+^ and IFN-γ^+^TNF-α^+^ within CD56^+^CD16^+^ NK or CD56^+^CD3^+^ NKT cells positively correlated with each other (table S4), demonstrating synchronized activation of all innate cells by DENV. In our large cohort, non-structural (NS) protein 1 levels, a measure of viral load did not vary with disease duration (fig. S10A). Patients with highest NS1 levels displayed highest innate cell cytokine secretion (fig. S10, B-D) and also avoided severe outcomes (fig. S10E), suggesting a requirement for high viral antigen levels to achieve efficient innate cell activation.

In order to query the link if any, between kinetics of innate immune activation by DENV and disease severity, we compared innate cell cytokine secretion between different measures of severity at early (days 1-3), intermediate (days 4-6) and late (days 7-15) times of hospital presentation. Patients admitted 1-3 days post symptom onset (dpso) had impressively higher TNF-α (Fig. 3A) and IFN-γ (Fig. 3B, S11A) -secreting innate cells in those with normal compared to above-normal liver enzyme levels. Those admitted 4-6 dpso had a significantly greater proportion of IFN-γ^+^CD56^+^CD3^+^ NKT cells as well as TNF-α-secreting granulocytes, CD19^+^ B cells and CD56^+^CD3^+^ NKT cells in DF relative to DFWS/SD (Fig. 3, C-F). In contrast, patients with high liver enzymes failed to down-regulate secretion of TNF-α, IL-6 and IP-10 from different innate cell subsets during the late stage (7-15 dpso) that was prominent in patients with normal levels (Fig. 3G, H & S11, B-D). Severe dengue was also characterized by a late surge of IP-10 (fig. S11, E-F). Dysregulated high levels of IFN-γ^+^CD56^+^CD3^+^ NKT cells and TNF-α^+^CD19^+^ B cells were also evident in DFWS/SD relative to DF patients during this late time window (fig. S11, G-H). Thus, robust early secretion of TNF-α and IFN-γ by innate cells during acute disease phase combined with efficient attenuation of all innate cytokines during later stages of disease averted severity.

**Fig. 3.**
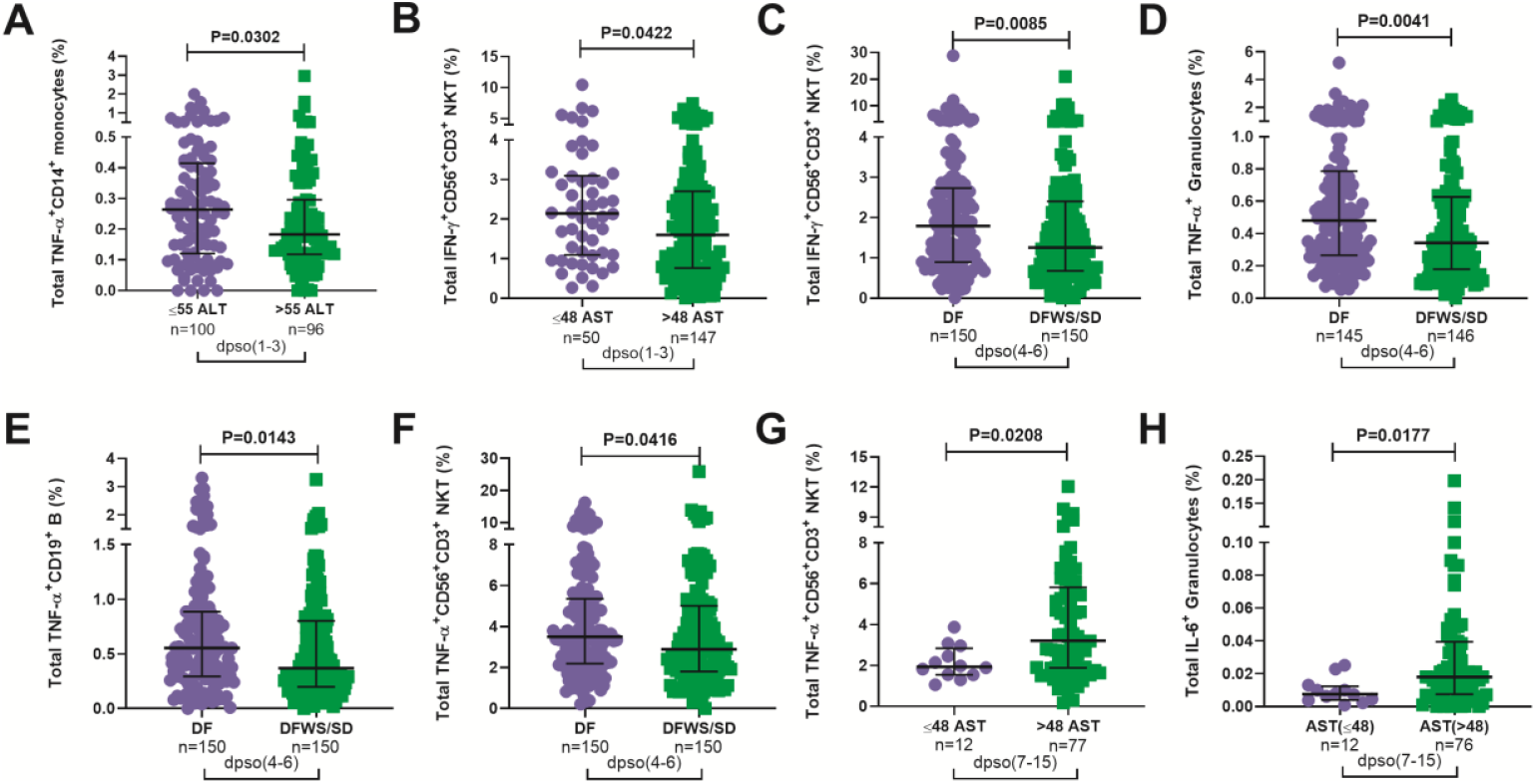
Correlation between disease severity and kinetics of innate immune cell activation. Frequency of total (**A**) TNF-α^+^CD14^+^ monocytes and (**B**) IFN-γ^+^CD56^+^CD3^+^ NKT cells during 1-3 dpso compared between normal and high levels of ALT (A; IU/L) and AST (B; IU/L). Frequency of total (**C**) IFN-γ^+^CD56^+^CD3^+^ NKT cells, (**D**) TNF-α^+^ granulocytes, (**E**) TNF-α^+^CD19^+^ B cells and (**F**) TNF-α^+^CD56^+^CD3^+^ NKT subsets during 4-6 dpso compared between DF and DFWS/SD. Frequency of total (**G**) TNF-α^+^CD56^+^CD3^+^ NKT cells and (**H**) IL-6^+^ granulocytes during 7-15 dpso compared between normal and high AST levels.

To identify potential biomarkers of progression to severity, we compared those who worsened after recruitment (as evidenced by a shift from DF/DFWS to DFWS/SD or death), with patients who readily recovered from DF and DFWS/SD. Both recovered DF and DFWS/SD patients carried a significantly greater proportion of IFN-γ^+^TNF-α^+^CD56^+^CD3^+^-, IFN-γ^+^CD56^+^CD3^+^-NKT cells and monofunctional IL-6^+^ granulocytes (Fig. 4, A-C) relative to worsened patients. To assess the biomarker performance of these cells, receiver operating characteristic (ROC) curve analysis was performed. IFN-γ^+^TNF-α^+^CD56^+^CD3^+^-, IFN-γ^+^CD56^+^CD3^+^ NKT and monofunctional IL-6^+^ granulocytes provided AUC of 0.77, 0.76 and 0.75 respectively with 90% sensitivity and 60 to 66% specificity when DF was compared with the worsened group (fig. S12, A-C). Combining IL-6^+^ granulocytes and IFN-γ^+^CD56^+^CD3^+^ NKT cells using binary logistic regression resulted in composite AUC of 0.85 (Fig. 4D; table S5) and revealed that every one percentage rise in IFN-γ^+^CD56^+^CD3^+^ NKT resulted in 3.12 fold lower odds of worsening (95% CI=1.1-9.2, P=0.035). In patients with elevated AST, this composite biomarker predicted the progression to severity with higher accuracy (AUC=0.9) and displayed 100% sensitivity with 81.9% specificity (table S5).

**Fig. 4.**
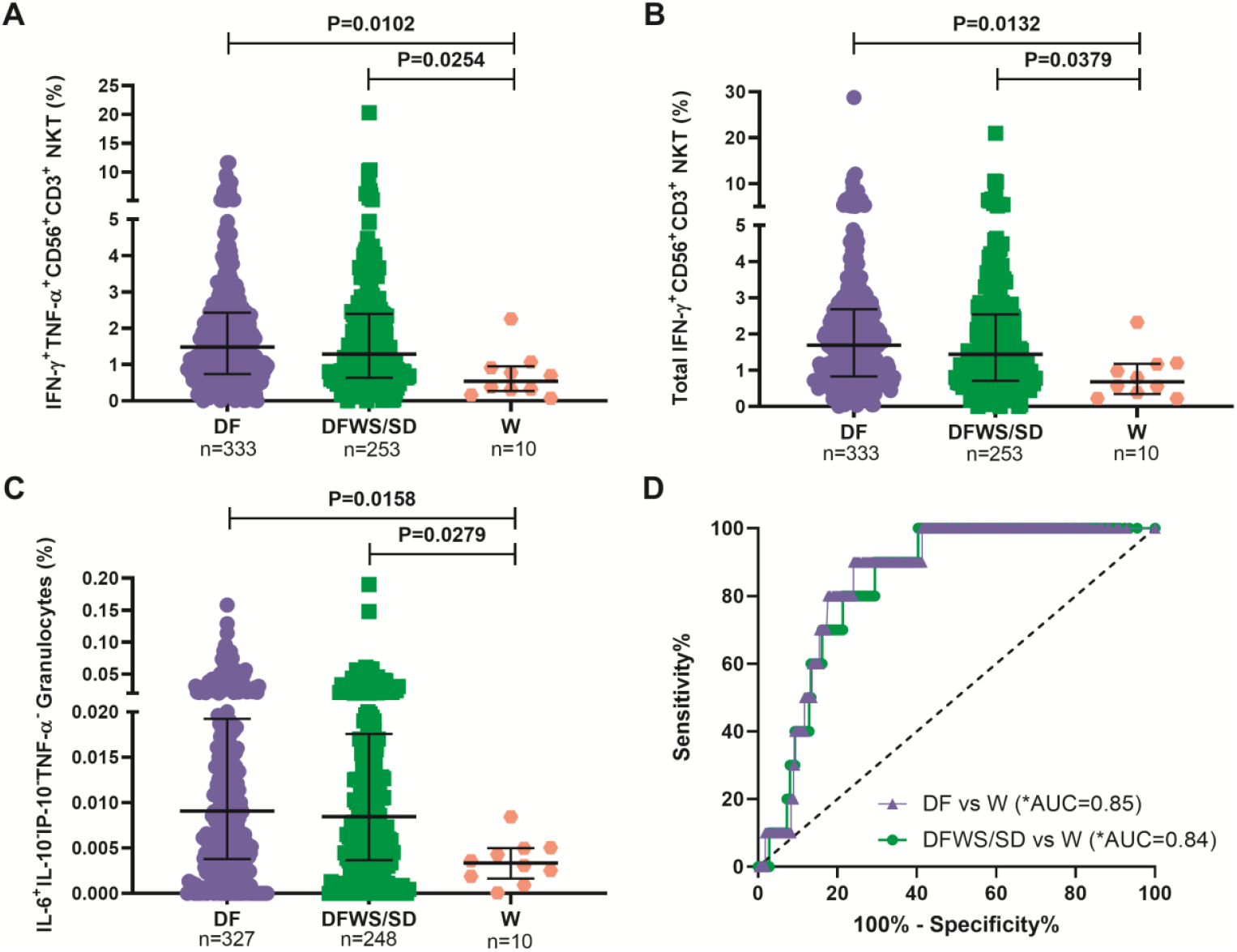
Innate cytokine secreting cells predicted outcome in dengue patients. Frequency of (**A**) IFN-γ^+^TNF-α^+^CD56^+^CD3^+^, (**B**) total IFN-γ^+^CD56^+^CD3^+^ NKT cells and (**C**) IL-6^+^IL-10^−^IP-10^−^TNF-α^-^ granulocytes compared between DF, DFWS/SD and worsened (W) patients. P value with median and IQR reported. (**D**) Composite ROC curve for total IFN-γ^+^CD56^+^CD3^+^ NKT cells and IL-6^+^IL-10^−^IP-10^−^TNF-α^-^ granulocytes comparing DF (purple) or DFWS/SD (green) with worsened patients. (*AUC), composite AUC.

This study, the first to query the cellular source of innate inflammatory cytokines in a large, blinded dengue cohort, conclusively demonstrated the beneficial role of early innate cell activation triggered by high viral antigen levels, in ensuring recovery from dengue. DENV activated all innate immune subsets resulting in mono and polyfunctional cytokine production that correlated with good outcome. The innate immune cytokine signature for each pathogen may be unique and rewarding to investigate.

The observed persistence of innate cell-derived cytokines during late phase of disease pointed to dysregulated innate responses in SD (*20*). Severe COVID-19 patients also displayed sustained plasma levels of IP-10 and IL-6 (*21*); increased plasma IP-10 also correlated with liver impairment in HIV/HBV patients (*22*). In light of the reported requirement of IP-10 for B cell activation (*23*), our finding of abnormal high IP-10 levels during late stages of disease is a likely contributor to the B cell mediated pathology attributed to severe dengue (*24*). A potential role for persistently elevated innate responses in provoking the reported inappropriate TCR signaling and T cell apoptosis in SD (*8, 9*) warrants further investigation. Thus, failure to achieve both robust activation and prompt attenuation of innate immune cells was a hallmark of severe dengue.

Our composite biomarker performed well despite limited number of patients with transitions to greater severity and inclusion of patients with varying disease duration. Potential enhancement of its performance by including additional hitherto unidentified cytokine secreting cells will enhance its utility in a clinical setting. Ease of processing and ready availability of flow cytometers in diagnostic laboratories assures feasibility of host blood-based biomarker deployment whereas biomarkers reliant on expensive instruments and high-end technical skills, may have limited utility in resource constrained geographies (*25, 26*).

CD56^+^CD16^+^ NK cells secreting TNF-α showed strong positive correlation with multiple subsets pointing to its central role in the coordinated innate immune activation by DENV. Our findings are in line with the 20 gene transcript signature that included anti-viral IFN-γ signaling pathway genes, being under-expressed in NK and NKT cells of SD patients (*26*). Weaker activation of early innate effector mechanisms was also reported in patients who later developed severe manifestations (*27*). The reported impaired innate immune activation in severe SARS-CoV-2 patients (*28*), suggests that severe disease in multiple viral infections may be a shared consequence of defective modulation of kinetics of early host innate activation as well as its subsequent attenuation.

## Data Availability

Data and materials availability: All data are available in the supplementary materials. De-identified flow cytometry files (.fcs) that support the results reported in this article, and analyzed flow cytometry data (.xlsx) will be made available on request.

## Acknowledgments

We sincerely thank all the study participants and the staff at BMCRI, RMCH, KIMS and SJMC. We thank Sivakami Sundari S for help with the interpretation of the logistic regression analysis, Madhusudan T for editing manuscript images and Chandrasekhar R for the safe transport of all clinical samples to Indian Institute of Science. We acknowledge Beckman Coulter - Bangalore Development Centre for providing the flow cytometer.

## Author contributions

VS conceived, designed, planned and supervised the study, interpreted data and wrote the manuscript. SPP and PHV conducted the experiments, analyzed data and wrote the manuscript. TT oversaw the statistical analysis of data. CR, MD, VK, YC, SD, NR, AVV, LS, RKB and MR were responsible for diagnosis, consenting and recruitment of patients, clinical sample collection and maintenance of case report forms.

## Data and materials availability

All data are available in the supplementary materials. De-identified flow cytometry files (.fcs) that support the results reported in this article, and analyzed flow cytometry data (.xlsx) will be made available on request.

## Supplementary Materials

Materials and Methods

Supplementary Text

Figures S1-S13

Tables S1-S11

References (*29, 30*)

STARD Checklist

Other Supplementary Materials for this manuscript include the following:

STARD Checklist

## Supplementary Materials for

### Materials and Methods

#### Ethics statement

This study was carried out in accordance with the Declaration of Helsinki. Institutional ethics committee approval to conduct the study was obtained from the four participating hospitals; Bangalore Medical College and Research Institute (BMCRI; BMCRI/PS/25/2018-19), Kempegowda Institute of Medical Sciences (KIMS; KIMS/IEC/A1-2018), St. John’s Medical College (SJMC; IEC/1/473/2019), M S Ramaiah Medical College (RMCH; MSRMC/EC/19) and Indian Institute of Science (IISc; 10-14032018), Bengaluru, India.

#### Study subjects and clinical data collection

778 participants were enrolled for this blinded study between June 24 and November 29, 2019 which represents the annual dengue season following onset of monsoon rains at the four hospitals listed above. Written informed consent was obtained from all participants before sample collection and analysis. Blood samples were coded and labeled as follows: DEN/hospital abbreviation/19/### before being sent to IISc for flow cytometry analysis. Consecutive suspected adult dengue patients (≥ 18 years) who tested positive using a dengue specific NS1/IgM rapid dengue day 1 test kit (J Mitra and Co., India) were recruited. Those who tested negative were recruited as febrile controls (FC); volunteers with no illness for the past 3 months were enrolled as healthy controls (HC; fig. S1).

Sample size analysis for a desired power of 80%, type I error tolerance of 0.05, and a hypothesized effect size of 0.75, required at least 29 dengue patients who would transition post admission, to a worse condition as defined by the World Health Organization (WHO) categorization of dengue severity as follows: dengue fever (DF) was defined by headache, body ache, rash, nausea, or mild bleeding; dengue fever with warning signs (DFWS) included symptoms like persistent vomiting, mucosal bleeding, pleural effusion, ascites, and hepatomegaly. Severe dengue (SD) included symptoms such as plasma leakage, ≥1000 IU/L of alanine aminotransferase (ALT) / aspartate aminotransferase (AST), severe bleeding which leads to shock, and/or organ impairment (*3*). However, when the dengue cases ceased to appear in the hospitals by end November 2019, we had obtained only 10 patients who transitioned to a worse category, primarily owing to the clinical interventions following admission. The study was also designed to collect longitudinal samples at days 3 and 7 post-admission; however, only 154 patients provided a second sample and a single patient donated three consecutive samples. Demographic characteristics (i.e., gender and age), clinical features (i.e., days post symptom onset, nausea, head ache, body ache, abdominal pain, rashes, splenomegaly, hepatomegaly and bleeding manifestations) and routine hematological laboratory findings (i.e., complete blood cell count, serum albumin, liver enzymes, platelet count and hematocrit) were recorded. Patients were assigned bleed-scores (BS) as follows: no bleeding, 0; petechiae, 1; epistaxis/gingival bleeding/menorrhagia, 2; gastrointestinal bleeding, 3; intracranial/intrapulmonary bleeding, 4. Plasma leakage in pleural and/or peritoneal cavities was confirmed using X-ray/ultrasound scans. HIV patients were not recruited. Data from samples of patients with co-infections (typhoid, sepsis, malaria, urinary tract infection, Hepatitis B) or those who were discharged against medical advice (DAMA) and samples with experimental errors (clotted blood samples, QC failure of flow cytometer, sample processing errors) were excluded from analysis.

#### Serology

Dengue virus (DENV) infection was confirmed using a commercial IgM, IgG and NS1 enzyme-linked immunosorbent assay (ELISA; Panbio, Australia) and results were interpreted according to manufacturer’s instructions. The kits were used to distinguish primary (IgM to IgG ratio >1.2) from secondary (IgM to IgG ratio <1.2) infection. Primary dengue status was also assigned to those who tested positive for DENV specific NS1 (index value >1.1) but were negative for IgM and IgG.

#### Ex vivo intracellular cytokine staining of innate immune cells in whole blood

Blood samples collected in sodium citrate vacutainer tubes (BD Biosciences) were immediately processed, no later than 4 hours from collection. RBCs from 500µl blood were lysed using 4ml of 1X ammonium chloride buffer (166mM ammonium chloride, 9.9mM potassium bicarbonate and 0.126mM EDTA). The centrifuged cells were washed with 1X phosphate buffered saline (PBS) and stained with Fixable Viability Stain 450 [BD, Cat#562247] for 10 minutes at room temperature, to exclude dead cells. This was followed by staining for appropriate surface markers (table S6) for 30 minutes at 4°C. The surface marker TLR2 was superior to HLA-DR owing to its stable expression during infection in contrast to the latter which is reported to be down-regulated in all manner of inflammatory conditions, and was therefore used to identify monocytes (*30*). Cells were fixed with 2% paraformaldehyde [Sigma Aldrich, Cat#P6148], washed and permeabilized with 0.1% saponin. The permeabilized cells were stained with intracellular antibodies (monocyte panel - IL-6, IP-10, IL-10, TNF-α; NK panel - IP-10, IL-10, TNF-α, IFN-γ) for 30 minutes at 4°C (table S6). Cells were washed, resuspended in 1X PBS and data were acquired on a Beckman Coulter DxFlex flow cytometer.

#### Analysis of flow cytometry data

Gating strategy for NK, monocyte and granulocyte subsets is shown in fig. S2. Cytokine-secreting cells are represented as percentage of parent population. Control samples were stained with surface antibodies and lacked all four intracellular markers keeping in mind the paucity of blood volumes (fluorescence minus four; FMF). Fluorescence minus one (FMO) controls were compared with FMF controls in five random individuals to confirm absence of non-specific binding of the intracellular cytokine-specific antibodies. FMF controls were used to set the positive gates for each cytokine (fig. S3). Positive cytokine production was based on the criteria given for each cytokine from each cell subset in tables S7 and S8. Data were analyzed using FlowJo software (version 10.6.1). Polyfunctional cytokine secretion was assessed by Boolean gating. The analyzed data from FlowJo was submitted to the clinical statistician for unblinding of patient characteristics prior to statistical analysis.

Optimized t-Distributed Stochastic Neighbor Embedding (t-SNE; (*31*)) analysis to visualize the clusters within the NK/NKT cells was performed. All 32 patient samples with SD were included along with 32 each from DF and DFWS (WHO 2009 categorization) which were selected at random using the RAND function in Excel 2016. A subset of 10,000 events were selected from CD19^-^ cells for each sample using DownSample plugin, followed by concatenation of all events. A total of 960,000 events and 7 markers (CD56, CD16, CD3, IP-10, IL-10, TNF-α, and IFN-γ) were used to generate the t-SNE map. We used the KNN algorithm (random projection forest – ANNOY) and Barnes-Hut gradient algorithm implementation with the recommended parameters (iterations – 1000; learning rate (eta) – 67200) at perplexity = 50 in t-SNE plugin built within FlowJo.

#### Statistical analysis

All analyses were done using IBM SPSS statistics 23.0 and GraphPad prism version 8. Significance between two or multiple groups was tested using Mann–Whitney *U* test (two-tailed) and non-parametric Kruskal-Wallis test with a Bonferroni correction for multiple comparisons, respectively. In patient cohort characteristics, normally distributed data were tested using one-way ANOVA. Chi square test of independence and Fisher’s exact test were used to evaluate the association of clinical parameters with WHO categorization of patients based on severity.

Differences between proportions of primary and secondary infection across WHO categories were assessed using the Z test for proportions. Confidence intervals for odds ratio were determined using Baptista-Pike method. Spearman’s correlation (two-tailed) analysis was performed to assess the positive or negative correlation between various cytokine secreting cell subsets. Receiver operating characteristic (ROC) curve analysis was performed to assess accuracy of proposed biomarker and 95% confidence intervals were calculated using Wilson/Brown method.

Multivariate binary logistic regression was performed to compare DF or DFWS/SD with worsened groups as the dependent variables. Independent variables for multivariate analysis were selected if they were significantly different in univariate analysis (two-tailed Mann–Whitney *U* test for non-parametric continuous data and Chi square test for categorical variables). Required assumptions such as dichotomous mutually exclusive dependent variable, two or more independent variables, linear relationship between each independent variable and odds ratio, absence of multicollinearity were all met. Even though three independent variables were significantly different two of them directly correlated with each other (IFN-γ^+^TNF-α^+^CD56^+^CD3^+^ NKT cells and total IFN-γ^+^CD56^+^CD3^+^ NKT cells). Hence we used a combination of monofunctional IL-6^+^ granulocytes with either total IFN-γ^+^CD56^+^CD3^+^ NKT cells or IFN-γ^+^TNF-α^+^CD56^+^CD3^+^ NKT cells to generate logistic regression models. The latter was not significant and was not used. Monofunctional IL-6^+^ granulocytes with total IFN-γ^+^CD56^+^CD3^+^ NKT cells regression model was a good fit confirmed by the Hosmer and Lemeshow goodness of fit test. The estimated probabilities obtained from logistic regression model were used to plot composite ROC curves.

## Supplementary Text

### Cohort Characteristics

Of the 596 subjects with laboratory confirmed dengue who were included in the final data analyses, the mean age was 30.4±10.79 (mean±SD, range 17-69). 72% of enrolled patients were male and 28% were female (table S9). They were admitted to the hospital at a median of 4 days (range 1-15) post symptom onset. Dengue specific IgM and IgG ELISA distinguished 336 (56.4%) primary patients from 256 (42.9%) secondary dengue patients. 281 patients tested positive for dengue specific NS1 ELISA. In our cohort, 333 (55.8%) patients were classified as DF, 227 (38.7%) as DFWS and 32 (5.4%) as SD. The clinical laboratory parameters that correlated with the diagnoses are listed in tables S10 and S11. The longitudinal samples collected were not used for data analysis/interpretation since the innate immune activation status in these samples reflected the effect of medical intervention rather than disease progression. We therefore used the day of presentation at hospital to query the alterations in immune status as a function of kinetics of disease progression. Post recruitment and hospitalization, 7 patients worsened sufficiently to transition from DF to DFWS and 1 from DFWS to SD while 2 died. Interestingly, 9 among 10 worsened patients were men.

### Cytokine signature of innate immune cells against DENV

The most abundant cytokines were IFN-γ from CD56^+^ NK cell subsets/CD19^+^ B cells and TNF-α from all innate cell subsets. CD16^+^ non-classical monocytes, CD56^+^CD3^+^ NKT cells, and CD14^+^CD16^+^ intermediate monocytes were the highest secretors of TNF-α. IFN-γ^+^TNF-α^+^ dual secreting cells were dominant in CD56^+^CD3^+^ NKT, CD19^+^ B cells, CD56^+^CD16^+^ and CD56^+^CD16^-^ NK cell subsets (fig. S12, A-D). CD56^+^CD3^+^ NKT cells also had abundant dual-functional IL-10^+^TNF-α^+^ cells followed by IP-10^+^TNF-α^+^ cells (fig. S12A). CD56^+^CD16^+^ NK cells carried polyfunctional profile dominated by IP-10 in combination with IFN-γ or TNF-α (fig. S12B). CD19^+^ B cells also showed the presence of IL-10^+^TNF-α^+^ dual functional cells followed by IP-10^+^TNF-α^+^ and IP-10^+^IFN-γ^+^. The triple positive cells from this subset included IFN-γ and TNF-α in combination with IL-10 or IP-10 (fig. S12C). Granulocytes and monocyte subsets had abundant IL-10^+^TNF-α^+^ cells (fig. S12, E-H). Granulocytes were the predominant secretor of multiple cytokines (IL-6, IL-10, IP-10 and TNF-α; table S1) in addition to multiple combinations of polyfunctional cells (fig S12H). IL-6 and IL-10 were the least abundant cytokines secreted by all queried innate cell types against DENV.

**Fig. S1.**
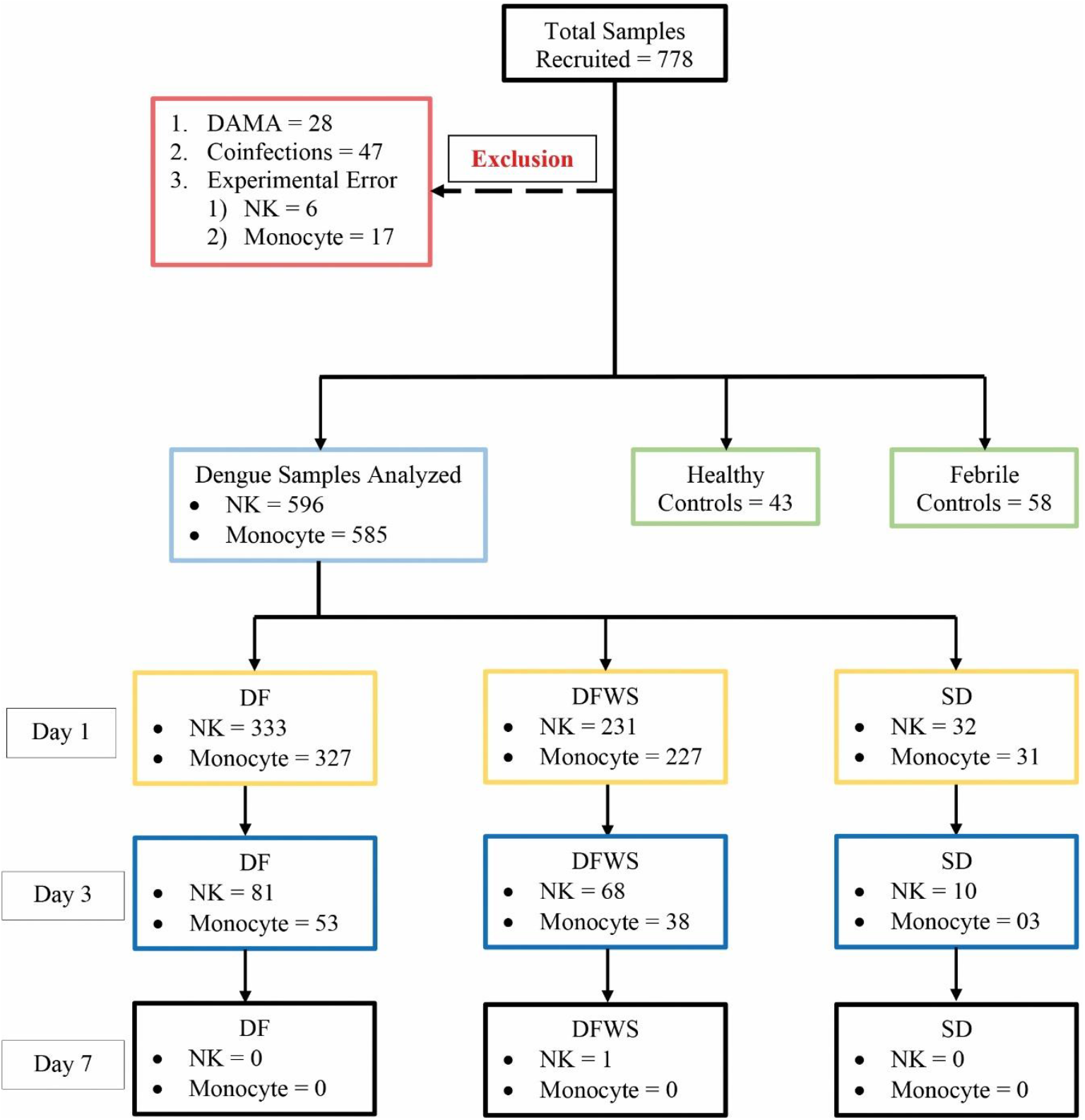
Schematic of patient recruitment. A total of 778 patients were initially recruited; patients who were discharged against medical advice (DAMA), those with co-infections (i.e. typhoid, sepsis, malaria, urinary tract infection, hepatitis B infection) and samples with experimental errors, were excluded as shown. Abbreviations: DF – dengue fever; DFWS – dengue fever with warning signs; SD –severe dengue, NK – natural killer cell.

**Fig. S2.**
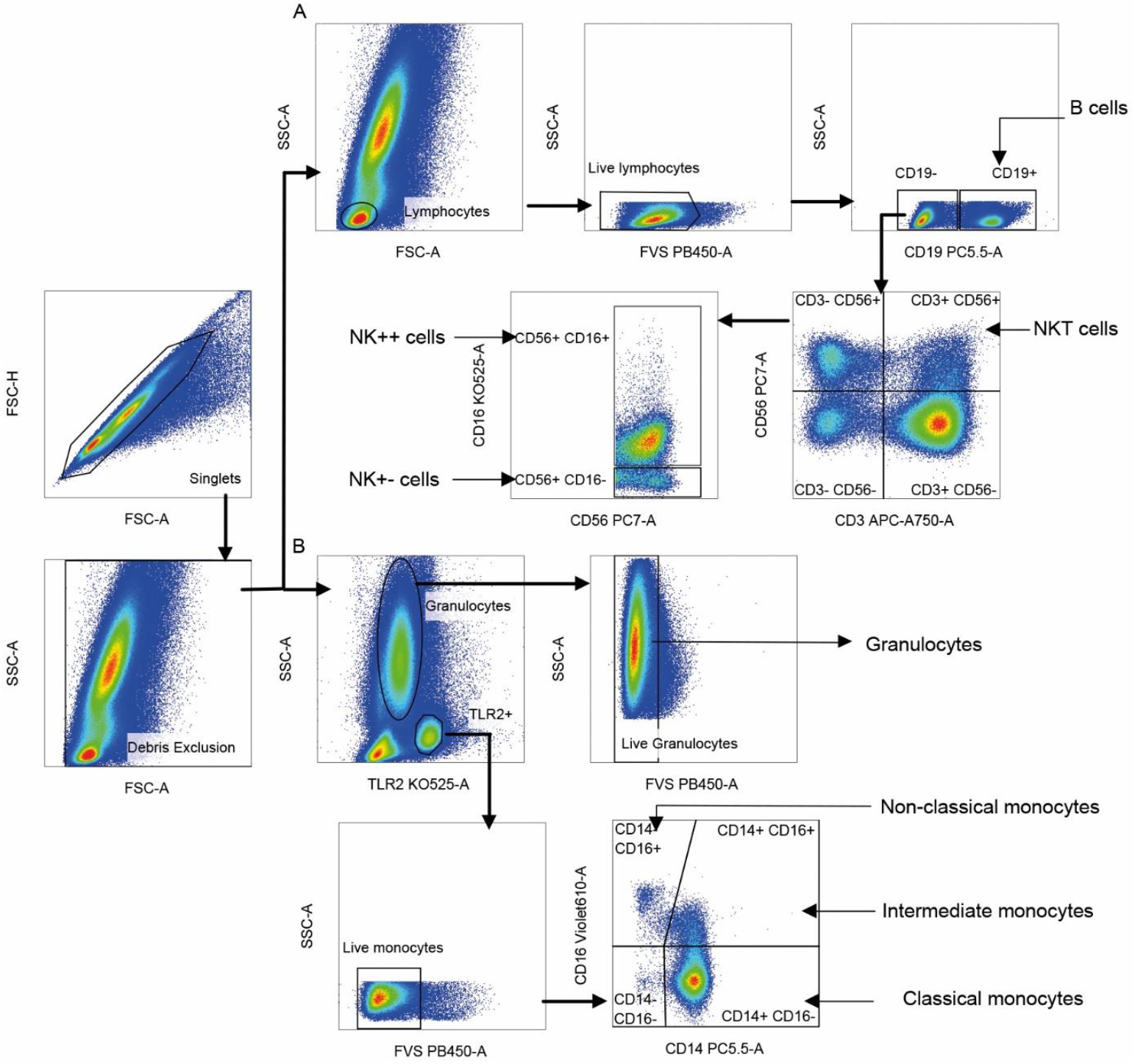
Gating strategy for innate immune cell subsets. Singlet cells were selected based on FSC-A and FSC-H scatter and debris was excluded based on FSC-A and SSC-A. (**A**) Lymphocytes were gated using FSC-A and SSC-A scatter; dead cells and B cells staining for live/dead dye and CD19, respectively were sequentially excluded. Natural Killer T (NKT) cells were identified as CD56^+^ CD3^+^ (CD56 vs. CD3). Two CD3-negative natural killer (NK) cells subsets were identified as CD56^+^ CD16^+^ (NK++) cells and CD56^+^ CD16^-^ (NK+-) cells displayed on CD56 vs. CD16. Cells positive for CD19 lineage marker were identified as B cells. (**B**) Live monocytes were identified as TLR2^+^ and negative for live/dead dye. Three monocyte subsets were distinguished as CD14^+^CD16^-^ classical monocytes (CM), CD14^+^CD16^+^ intermediate monocytes (IM) and CD14^-^ CD16^+^ non-classical monocytes (NCM) based on CD16 vs. CD14. Live granulocytes were distinguished based on SSC-A scatter vs TLR-2 followed by those negative for live/dead dye.

**Fig. S3.**
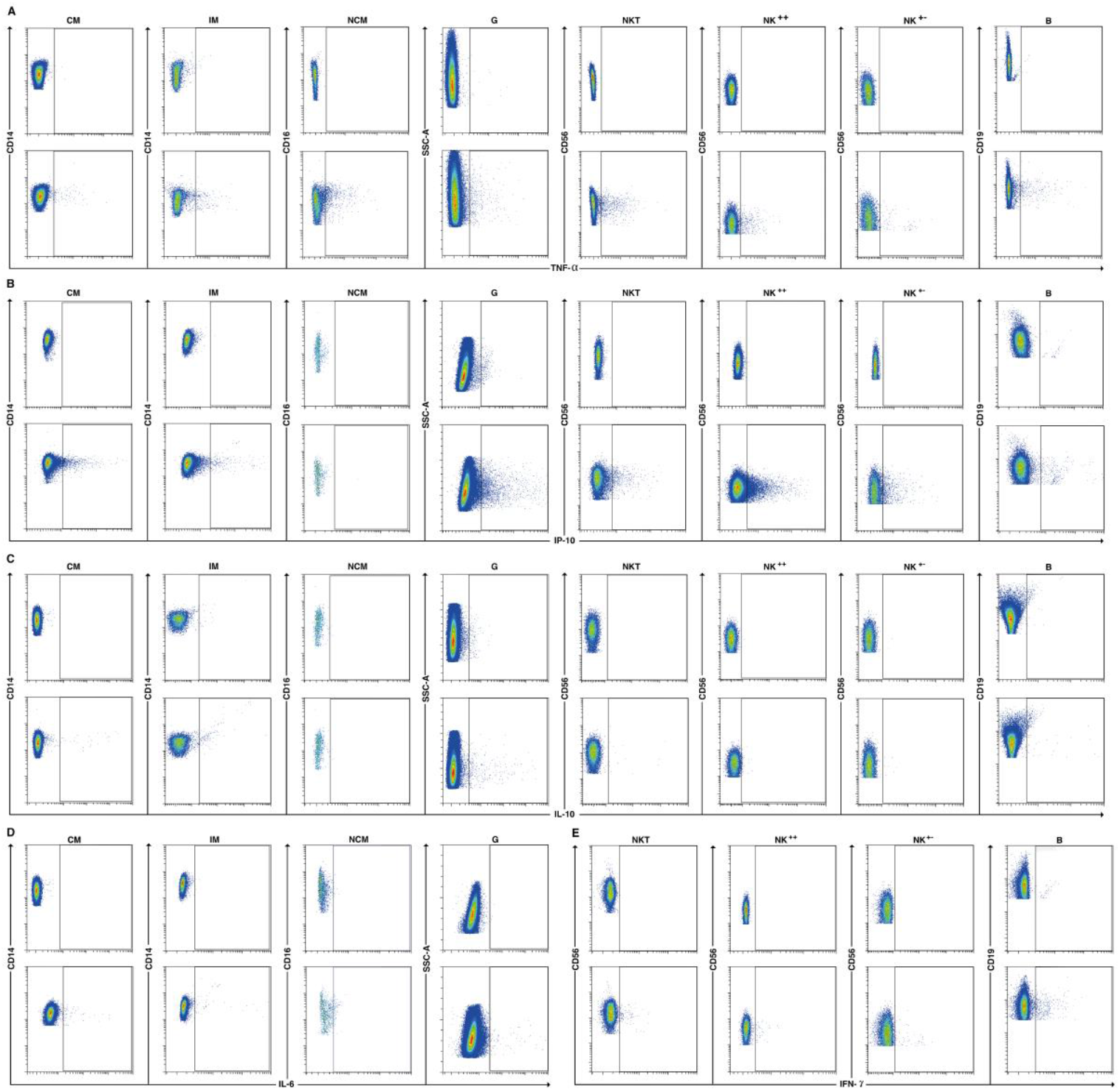
Fluorescence minus four controls for flow cytometry. Pseudo color flow cytometry plots for a representative patient comparing the fluorescence minus four (FMF; top row) control with completely stained sample (bottom) for secretion of (**A**) TNF-α, (**B**) IP-10, (**C**) IL-10, (**D**) IL-6 and (**E**) IFN-γ from the indicated cell subsets. Abbreviations: CM – CD14^+^CD16^-^ classical monocytes; IM – CD14^+^CD16^+^ intermediate monocytes; NCM – CD14^-^CD16^+^ non-classical monocytes; G – Granulocytes; NKT – CD56^+^CD3^+^ cells; NK^++^ - CD56^+^CD16^+^ cells; NK^+-^ - CD56^+^CD16^-^ cells; B – CD19^+^ cells.

**Fig. S4.**
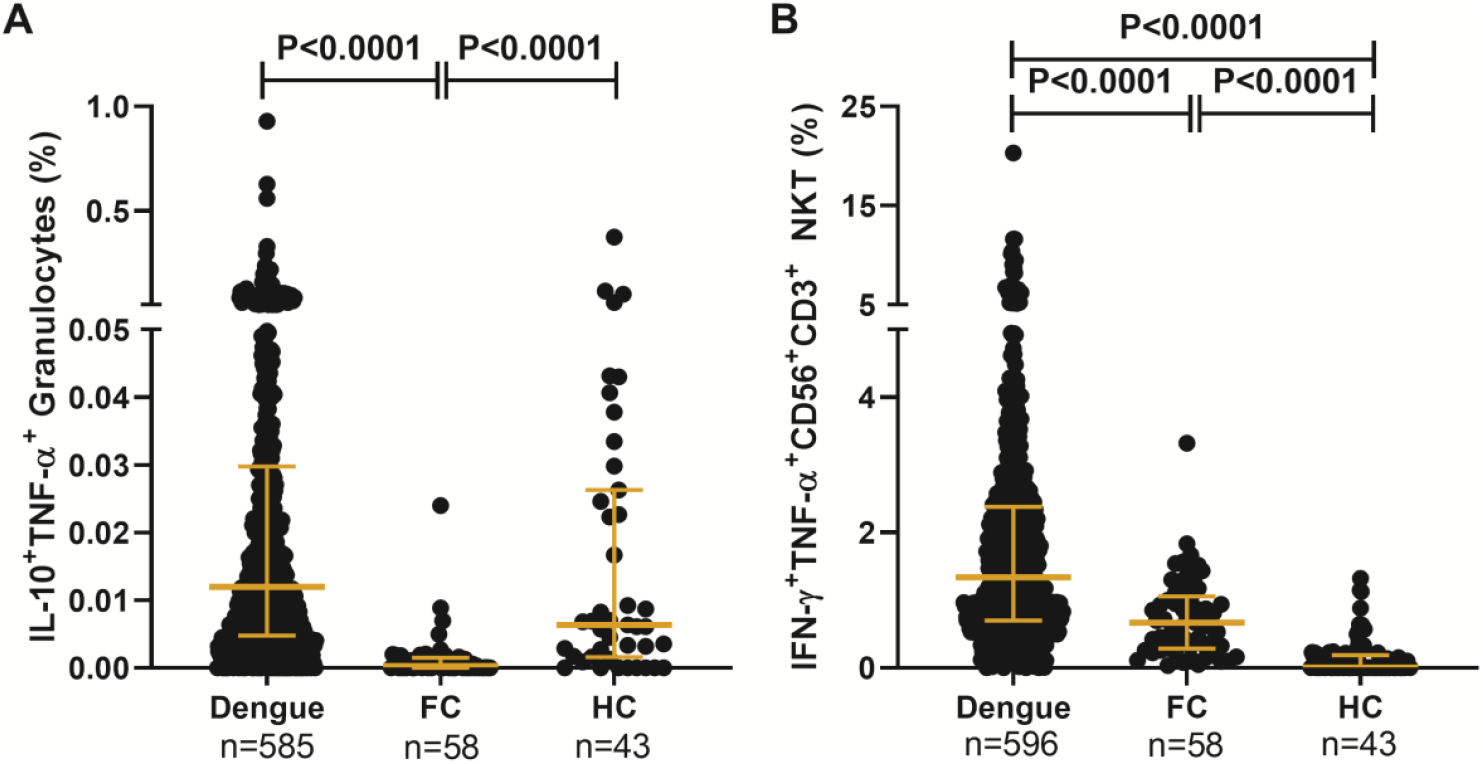
DENV activates dual-functional cytokine secreting innate immune cells. Frequency of (**A**) IL-10^+^TNF-α^+^ granulocytes and (**B**) IFN-γ^+^TNF-α^+^CD56^+^CD3^+^ NKT cells compared between dengue patients, febrile controls (FC) and healthy controls (HC). Each dot represents a patient sample. P value determined using Kruskal-Wallis test, followed by Bonferroni correction for multiple comparisons with median and IQR reported.

**Fig. S5.**
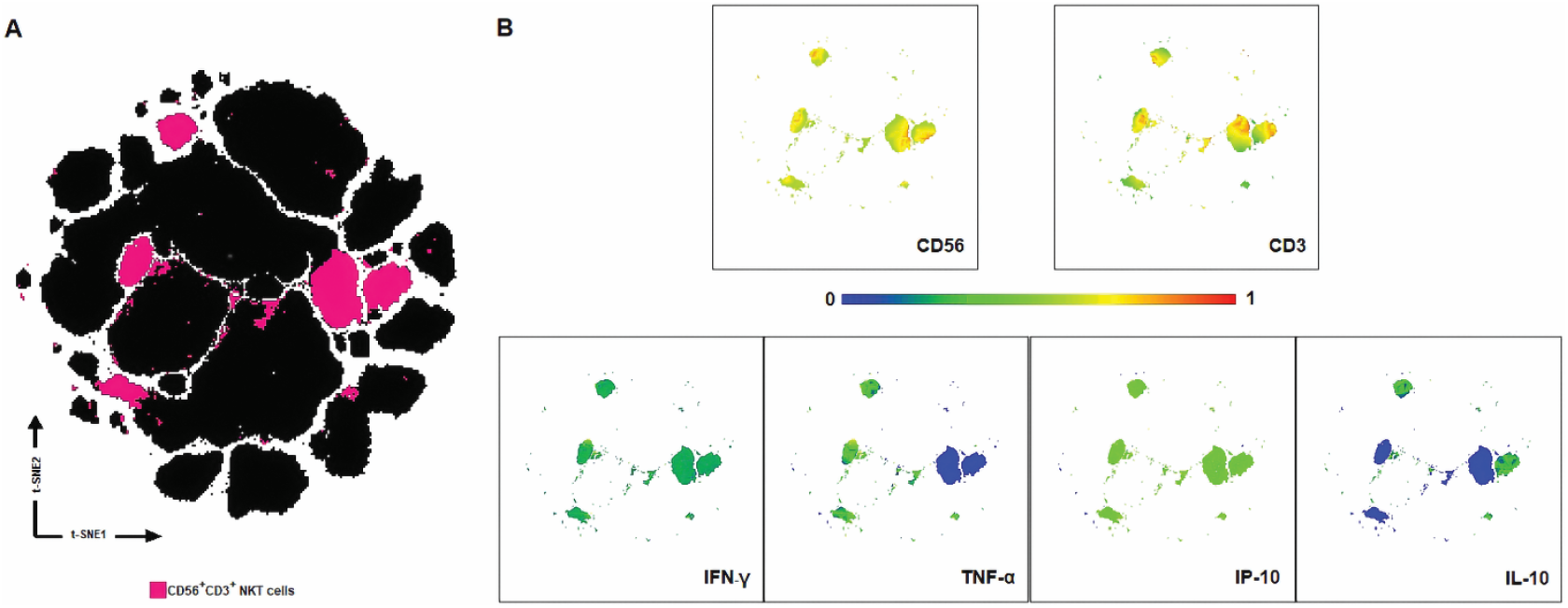
Visualization of CD56^+^CD3^+^ NKT cells in t-SNE dimensional reduced space. **(A)** Two-dimensional representation identified five major CD56^+^CD3^+^ NKT cell clusters (pink) visualized by t-SNE map. **(B)** Differential expression of CD56, CD3, IFN-γ, TNF-α, IP-10 and IL-10 in NKT cell clusters visualized by two-dimensional multicolored t-SNE maps.

**Fig. S6.**
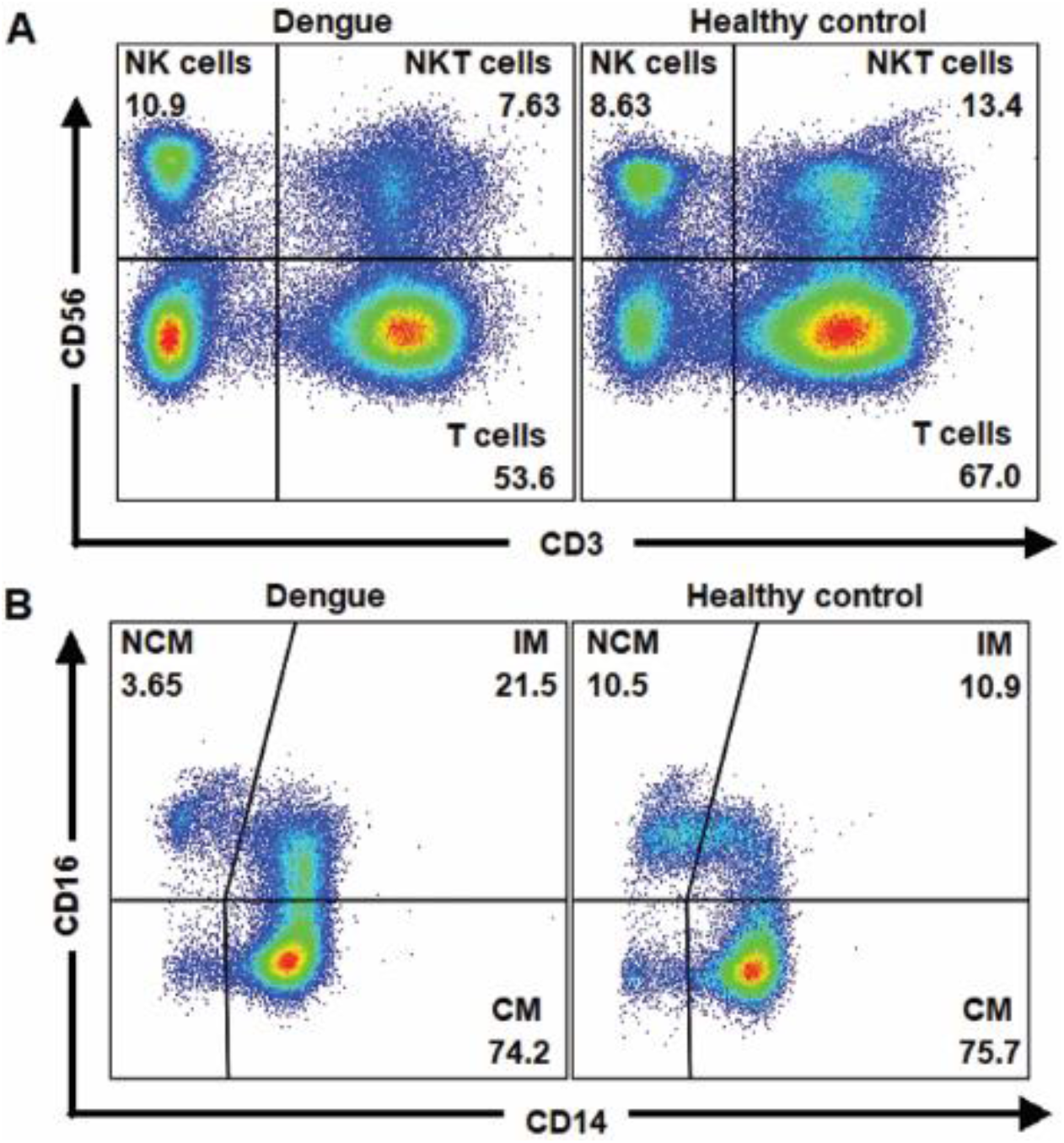
Modulation of innate immune cell subsets by dengue. Pseudo color flow cytometry plots for a representative patient and healthy control show (**A**) reduction in percentages of CD56^+^CD3^+^ NKT cells and (**B)** expansion of percentage of CD14^+^CD16^+^ intermediate monocytes in dengue patients relative to healthy control. Abbreviations: NK - CD56^+^ cells; NKT – CD56^+^CD3^+^ cells; T – CD3^+^ T cells; NCM – CD14^-^CD16^+^ non-classical monocytes; IM – CD14^+^CD16^+^ intermediate monocytes; CM – CD14^+^CD16^-^ classical monocytes.

**Fig. S7.**
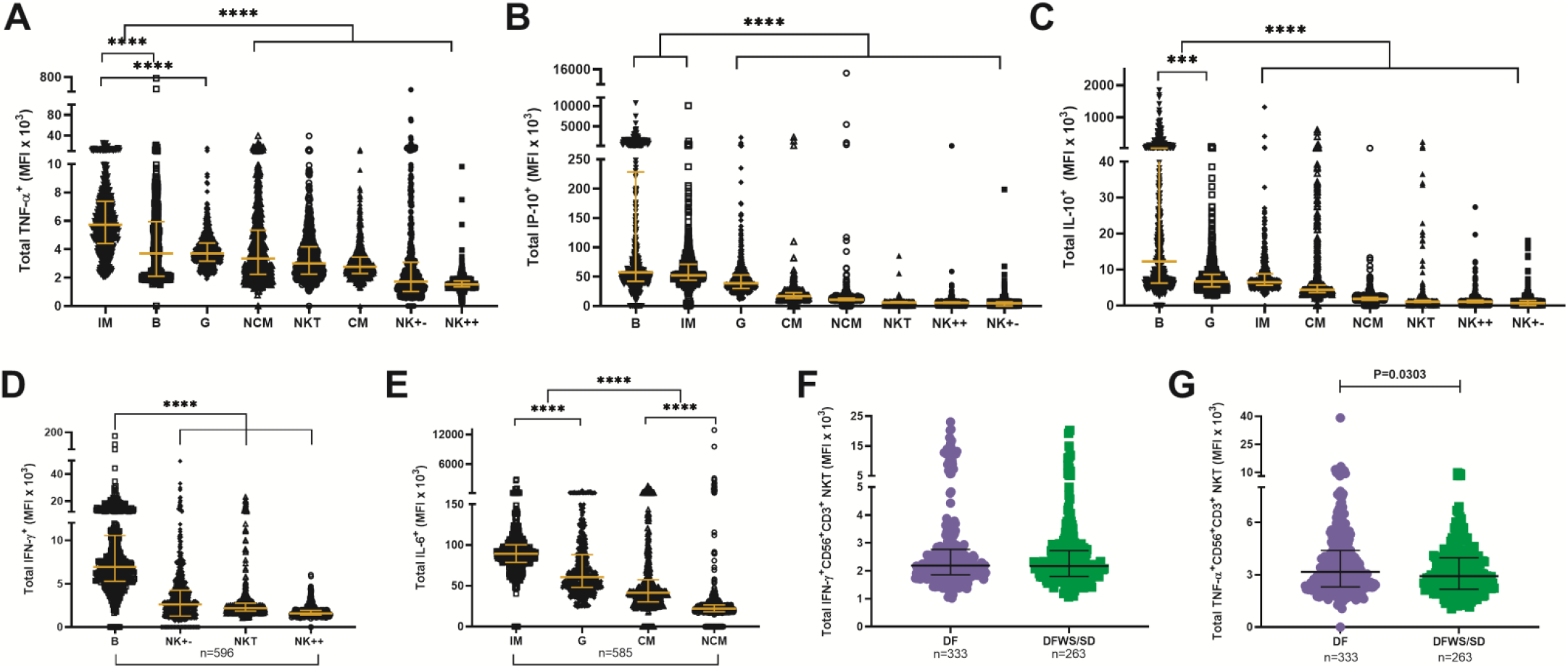
Per cell secretion of cytokines from each cell subset. MFI values for each cytokine were compared between subsets. (**A**) TNF-α (**B**) IP-10 (**C**) IL-10 (**D**) IFN-γ (**E**) IL-6 compared among cell subsets. P values were determined using Kruskal-Wallis test, followed by Bonferroni correction for multiple comparison between groups with median and IQR reported. **** P<0.0001. MFI of (**F**) total IFN-γ^+^ and (**G**) total TNF-α^+^ cells within CD56^+^CD3^+^ NKT cells compared between DF and DFWS/SD. Mann-Whitney *U* test was performed and median with IQR are reported. Abbreviations: MFI – median florescence intensity; CM – CD14^+^CD16^-^ classical monocytes; IM – CD14^+^CD16^+^ intermediate monocytes; NCM – CD14^-^CD16^+^ non-classical monocytes; G – Granulocytes; NKT – CD56^+^CD3^+^ cells; NK^++^ - CD56^+^CD16^+^ cells; NK^+-^ - CD56^+^CD16^-^ cells; B – CD19^+^ B cells.

**Fig. S8.**
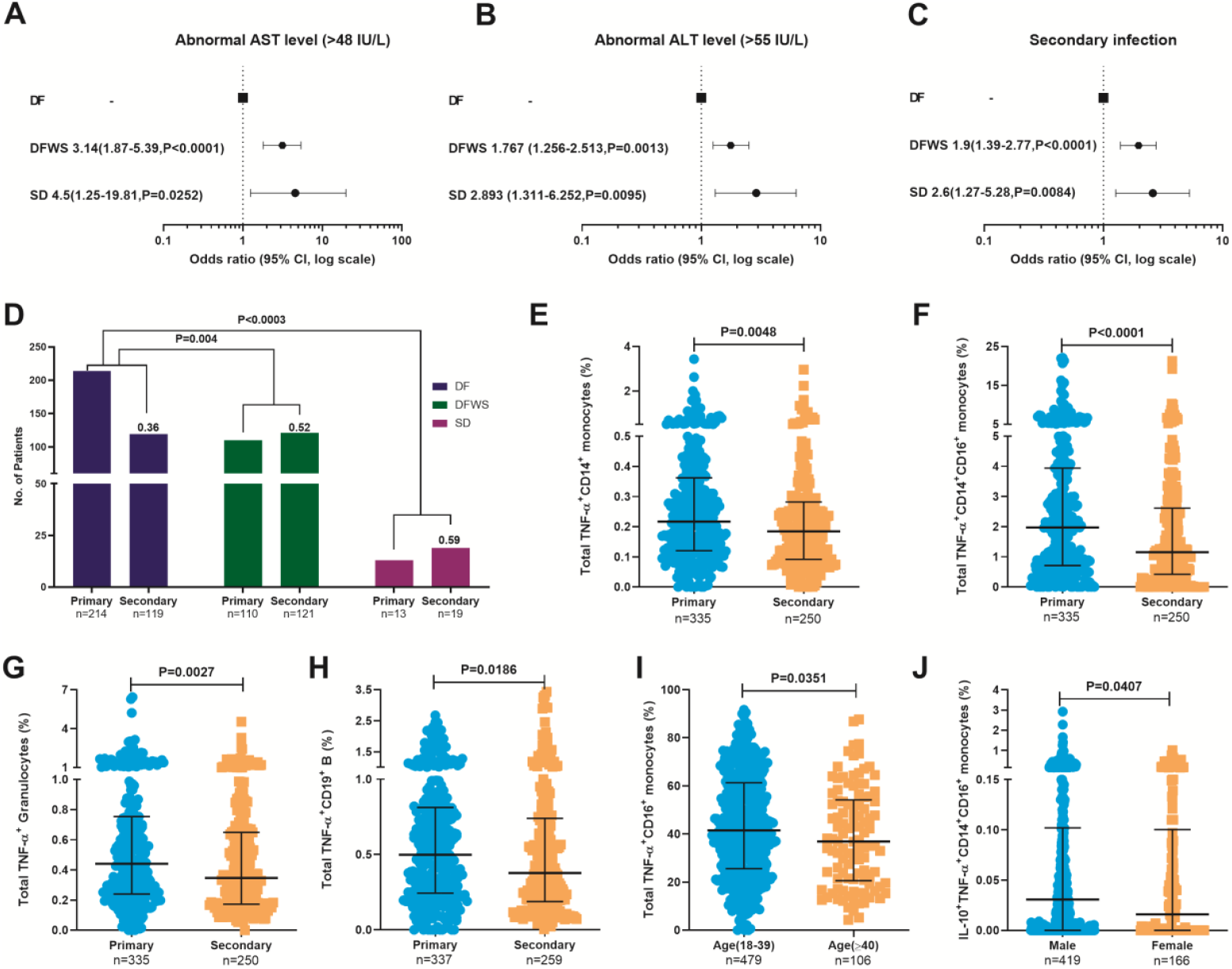
Factors influencing innate immune cytokine secretion in dengue patients. Forest plot showing association of dengue severity with (**A**) AST levels, (**B**) ALT levels and (**C**) serostatus. Data are odds ratio (95% CI, P value). (**D**) Z-score test to compare the proportion of primary/secondary dengue in DF, DFWS and SD. Significantly higher frequency of total (**E**) TNF-α^+^CD14^+^ monocytes, (**F**) TNF-α^+^CD14^+^CD16^+^ intermediate monocytes, (**G**) TNF-α^+^ granulocytes and (**H**) TNF-α^+^CD19^+^ B cells in primary compared to secondary patients. (**I**) Significantly higher frequency of total TNF-α^+^CD16^+^ non-classical monocytes in young (18-39 years) compared to old (≥40 years) patients. (**J**) Significantly higher frequency of IL-10^+^TNF-α^+^ CD14^+^CD16^+^ intermediate monocytes in men compared to women. P values were determined using Mann-Whitney *U* test and median with IQR are reported.

**Fig. S9.**
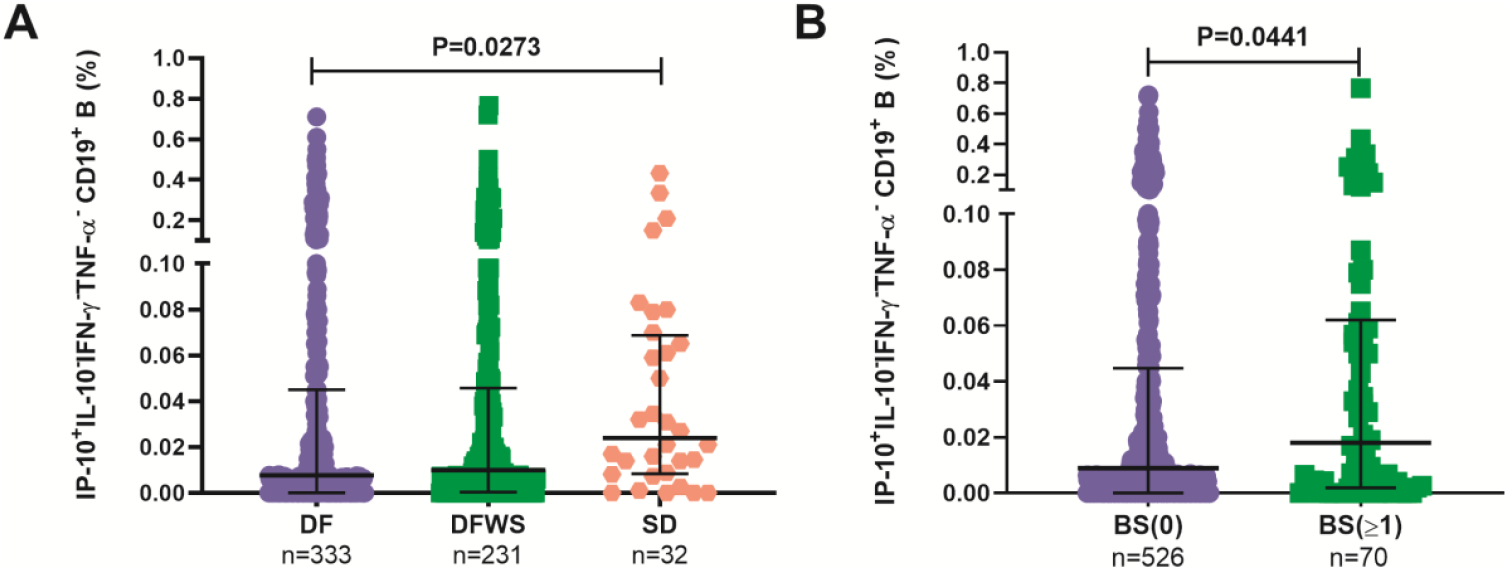
IP-10 secreting CD19^+^ B cells directly correlate with severity. Frequency of monofunctional IP-10^+^IL-10^−^IFN-γ^-^TNF-α^-^CD19^+^ B cells were **(A)** significantly higher in SD compared to DF and **(B)** significantly higher in patients with different degrees of hemorrhage (BS≥1) compared to patients with no bleeding (BS=0). P values were determined using Kruskal-Wallis test, followed by Bonferroni correction for multiple comparisons and using Mann-Whitney *U* test between two groups with median and IQR reported.

**Fig. S10.**
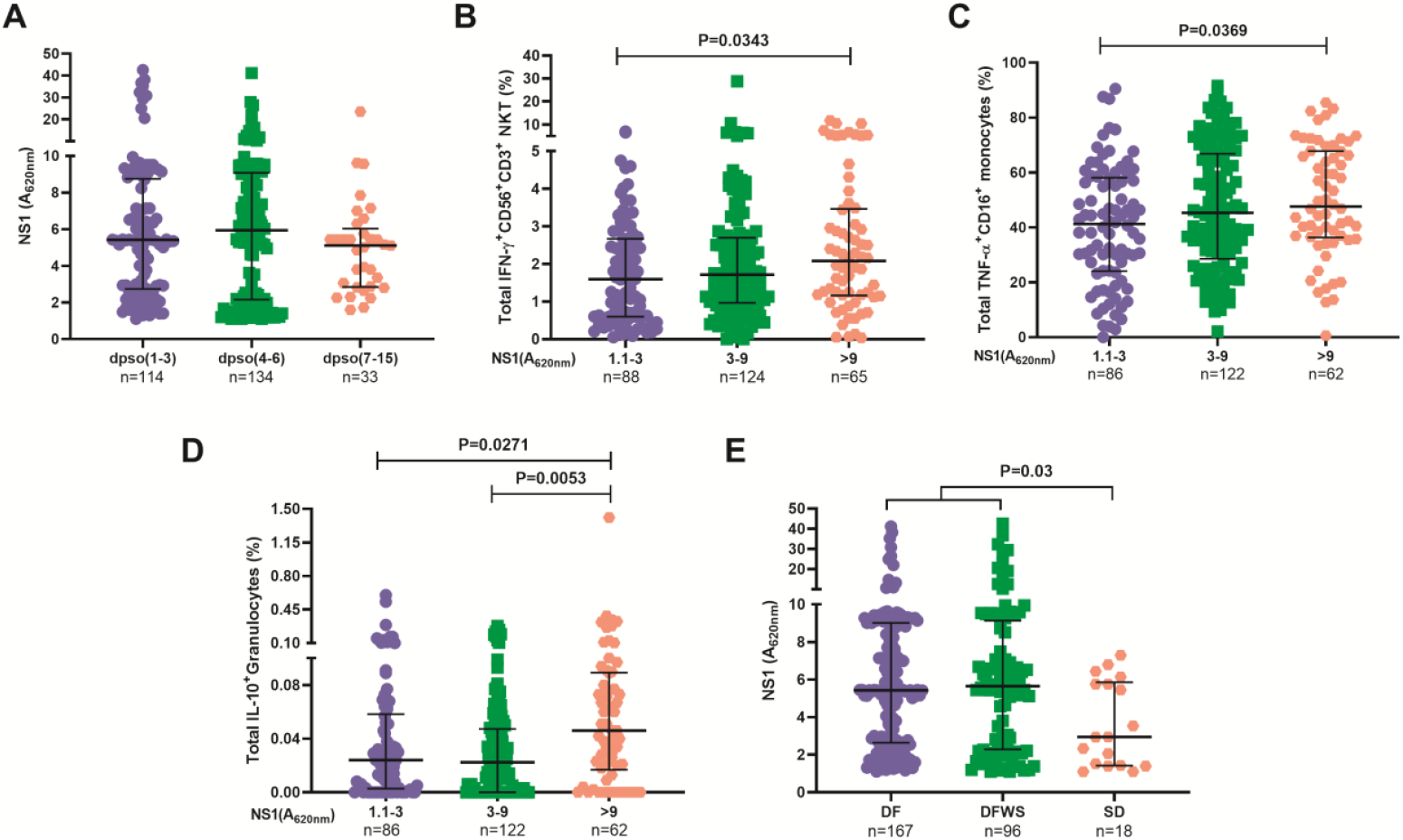
DENV NS1 protein levels correlate directly with innate immune activation. **(A)** All patients who had detectable dengue specific NS1 levels in serum were compared based on days post symptom onset (dpso). Frequency of total (**B**) IFN-γ^+^CD56^+^CD3^+^ NKT cells, (**C**) TNF-α^+^CD16^+^ non-classical monocytes and (**D**) IL-10^+^ granulocytes compared between patients with low NS1 (A_620nm_ 1.1 to 3), intermediate NS1 (A_620nm_ 3 to 9) and high NS1 (A_620nm_ > 9) values. (**E**) Levels of dengue NS1 compared between DF, DFWS and SD patients. P values were determined using Kruskal-Wallis test, followed by Bonferroni correction for multiple comparison between groups with median and IQR reported.

**Fig. S11.**
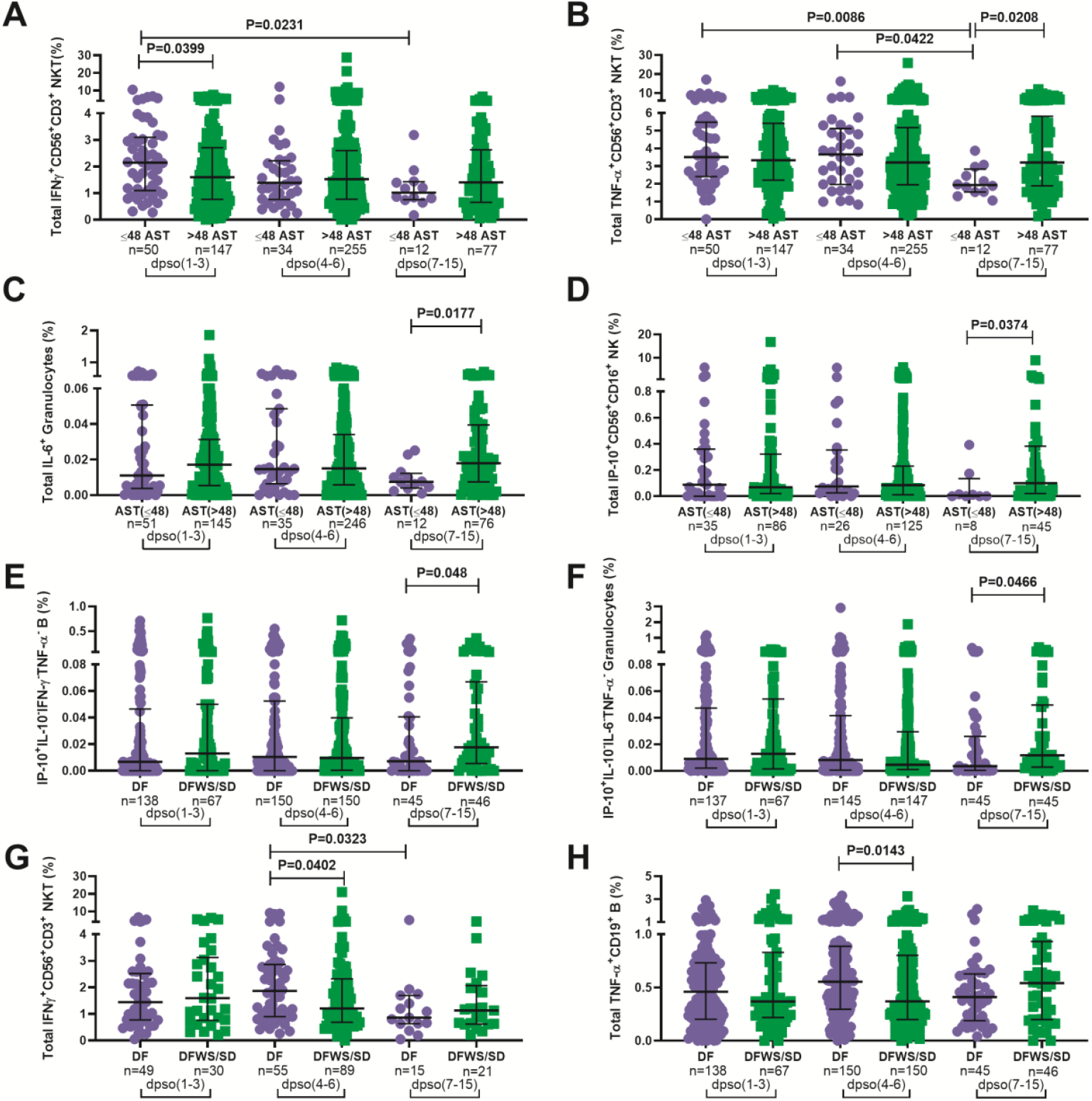
Kinetics of cytokine secretion by innate immune cells in dengue patients. Frequency of total (**A**) IFN-γ^+^CD56^+^CD3^+^ NKT cells, (**B**) TNF-α^+^CD56^+^CD3^+^ NKT cells, (**C**) IL-6^+^ granulocytes in total cohort and (**D**) IP-10^+^CD56^+^CD16^+^ NK cells in primary dengue cohort compared between normal (≤48 IU/L) and abnormal (>48 IU/L) levels of AST. Frequency of (**E**) monofunctional IP-10^+^IL-10^−^IFN-γ^-^TNF-α^-^CD19^+^ B cells in total cohort, (**F**) monofunctional IP-10^+^IL-10^−^IL-6^-^TNF-α^-^ granulocytes in total cohort, (**G**) total IFN-γ^+^CD56^+^CD3^+^ NKT cells in secondary dengue and (**H**) total TNF-α^+^CD19^+^ B cells in total cohort compared between DF and DFWS/SD severity groups as well as between time window of hospital presentation during 1-3, 4-6 or 7-15 days post symptom onset (dpso). P values were determined using Mann-Whitney *U* test between normal and abnormal levels of AST or DF and DFWS/SD groups for any single time interval and Kruskal-Wallis test, followed by Bonferroni correction for multiple comparison of normal or abnormal AST / DF or DFWS/SD patients between the three time intervals. Medians with IQR are reported.

**Fig. S12.**
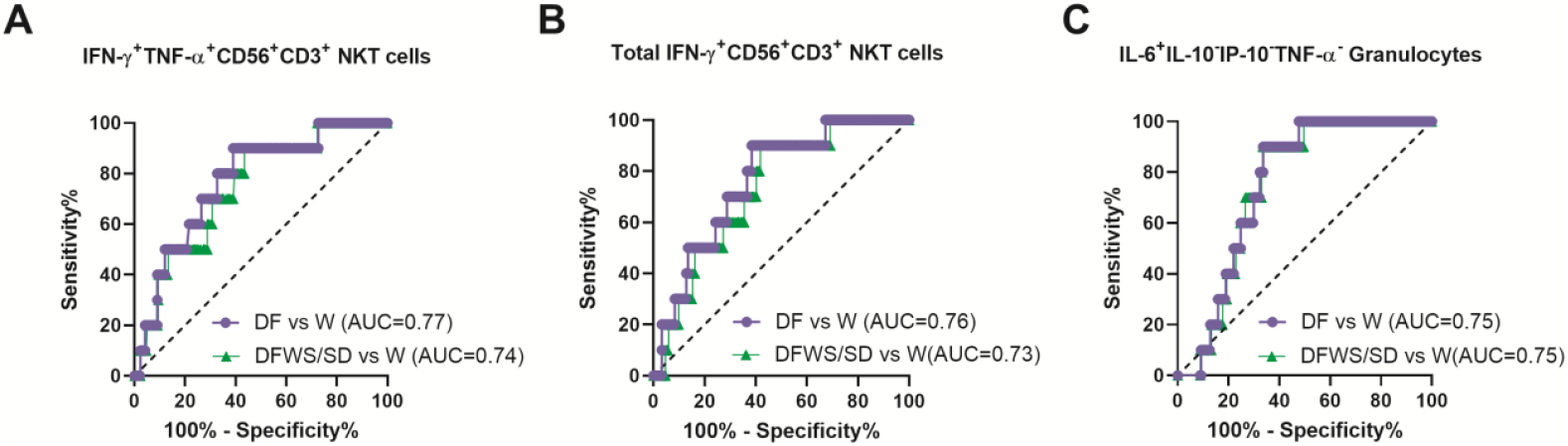
Biomarker performance of cytokine-secreting innate immune cell subsets. Receiver operating characteristic curve of **(A)** IFN-γ^+^TNF-α^+^CD56^+^CD3^+^ NKT cells, **(B)** total IFN-γ^+^CD56^+^CD3^+^ NKT cells and **(C)** monofunctional IL-6^+^IL-10^−^IP-10^−^TNF-α^-^ granulocytes in discriminating worsened (W) patients from recovered DF (purple) or DFWS/SD (green) patients. Area under the curves (AUC) were determined by ROC curve analyses.

**Fig. S13.**
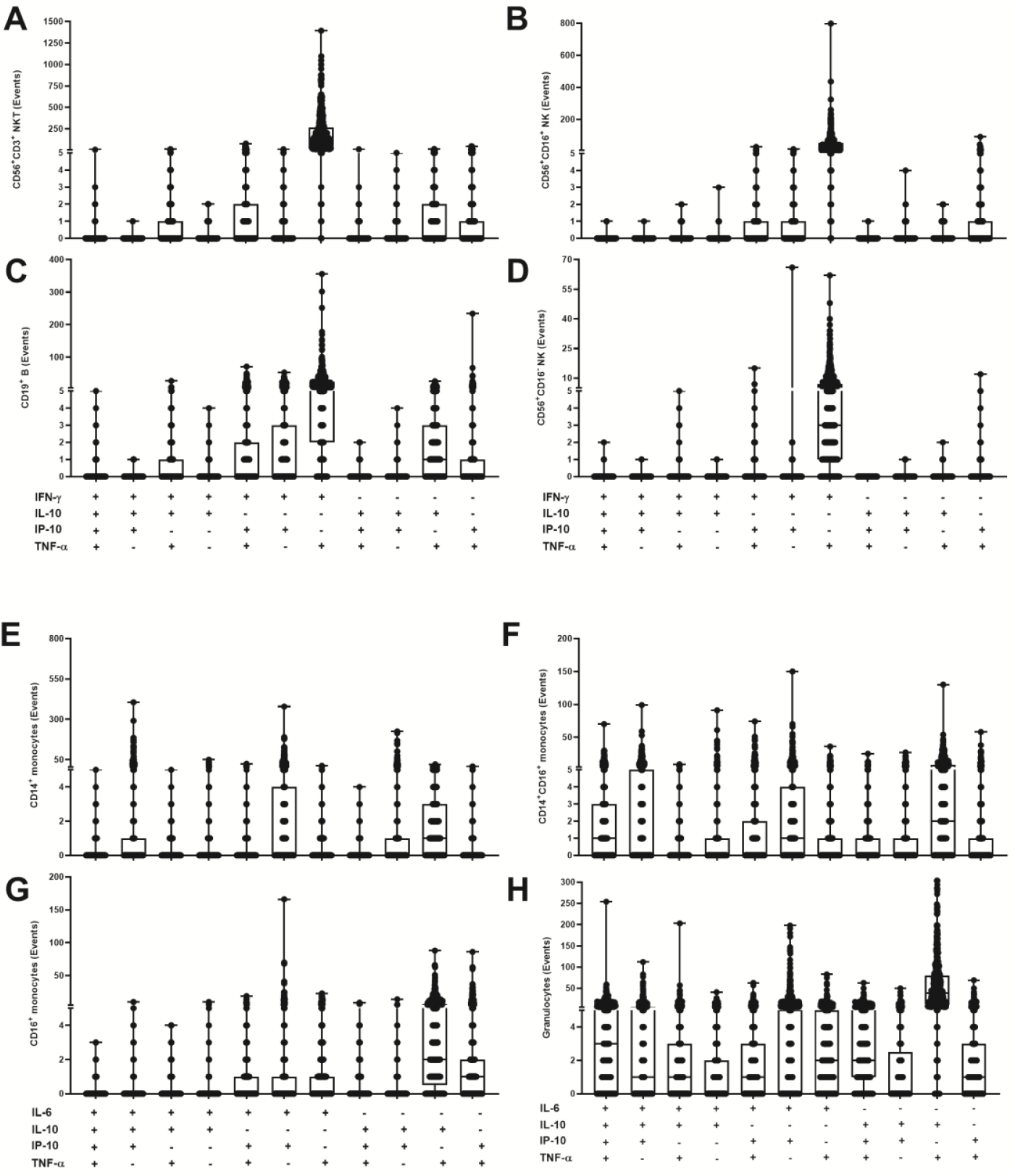
Polyfunctional profiles of innate immune cells. Polyfunctional cytokine signature of **(A)** CD56^+^CD3^+^ NKT cells **(B)** CD56^+^CD16^+^ NK cells **(C)** CD19^+^ B cells **(D)** CD56^+^CD16^-^ NK cells **(E)** CD14^+^ classical monocytes **(F)** CD14^+^CD16^+^ intermediate monocytes **(G)** CD16^+^ non-classical monocytes and **(H)** Granulocytes. Cytokine-secreting cell events over the control are plotted as box and whisker plot. Each dot represents a patient.

**Table S1.** Percentage producers and MFI of cytokines from innate cell subsets. % producers describe the percentage of patients in our cohort with detectable cytokine events from each cell subset; median cytokine secreting cells as a percent of parent and IQR are represented within brackets. The tabulated MFI (AU x 10^3^) are median (IQR). Abbreviations: MFI - Median Fluorescence Intensity; AU – Arbitrary Unit; IQR – Inter Quartile Range; NKT – CD56^+^CD3^+^ cells; NK^++^ - CD56^+^CD16^+^ cells; NK^+-^ - CD56^+^CD16^-^ cells; B cells – CD19^+^ cells; CM – CD14^+^ classical monocytes; IM – CD14^+^CD16^+^ intermediate monocytes; NCM – CD16^+^ non-classical monocytes.

| Cell Subsets | IFN-γ | | IP-10 | | IL-10 | | TNF-α | |
| --- | --- | --- | --- | --- | --- | --- | --- | --- |
|  | *% Producers*<br>*[Median (IQR)]* | *MFI*<br>*(AU x 10<sup>3</sup>)* | *% Producers*<br>*[Median (IQR)]* | *MFI*<br>*(AU x 10<sup>3</sup>)* | *% Producers*<br>*[Median (IQR)]* | *MFI*<br>*(AU x 10<sup>3</sup>)* | *% Producers*<br>*[Median (IQR)]* | *MFI*<br>*(AU x 10<sup>3</sup>)* |
| **NKT** | 95.5<br>[1.5 (0.77-2.6)] | 2.2<br>(1.8-2.7) | 44.3<br>[0.06 (0.004-0.3)] | 6.1<br>(5.4-7.4) | 33.9<br>[0.04 (0.007-0.1)] | 1.1<br>(0.8-1.5) | 98.2<br>[3.25 (2.0 -5.3)] | 3.0<br>(2.3-4.2) |
| **NK++** | 81.9<br>[0.29 (0.16-0.56)] | 1.6<br>(1.3-1.9) | 48.3<br>[0.065(0.1-0.28)] | 5.5<br>(4.6-6.9) | 13.6<br>[0.009 (0-0.03)] | 1.0<br>(0.8-1.5) | 95.1<br>[1.007 (0.57-1.61)] | 1.5<br>(1.3-1.8) |
| **NK+-** | 13.8<br>[0.13 (0-0.33)] | 2.6<br>(1.3-4.2) | 21.0<br>[0.29 (0-0.29)] | 4.8<br>(0-6.9) | 1.5<br>[0 (0-0.03)] | 0.7<br>(0-1.4) | 44.1<br>[0.52 (0.2-0.9)] | 1.7<br>(1.0-3.1) |
| **B cell** | 50.8<br>[0.07 (0.02-0.16)] | 6.9<br>(5.3-10.6) | 38.3<br>[0.02 (0.002-0.049)] | 57.3<br>(42.1-228.6) | 27.7<br>[0.017 (0.003-0.049)] | 12.2<br>(6.2-41.2) | 88.6<br>[0.44 (0.22-0.78)] | 3.7<br>(2.1-5.9) |
| | IL-6 | | IP-10 | | IL-10 | | TNF-α | |
|  | *% Producers*<br>*[Median (IQR)]* | *MFI*<br>*(AU x 10<sup>3</sup>)* | *% Producers*<br>*[Median (IQR)]* | *MFI*<br>*(AU x 10<sup>3</sup>)* | *% Producers*<br>*[Median (IQR)]* | *MFI*<br>*(AU x 10<sup>3</sup>)* | *% Producers*<br>*[Median (IQR)]* | *MFI*<br>*(AU x 10<sup>3</sup>)* |
| **CM** | 28.2<br>[0.01 (0-0.14)] | 41.6<br>(30.3-57.4) | 47.0<br>[0.027 (0-0.13)] | 16.9<br>(12.9-22.9) | 35.6<br>[0.01 (0-0.05)] | 4.4<br>(3.6-5.7) | 87.9<br>[0.2 (0.11-0.32)] | 2.8<br>(2.3-3.5) |
| **IM** | 37.8<br>[0.31 (0-0.2)] | 89.2<br>(78.8-100.3) | 41.2<br>[0.05 (0-0.25)] | 52.2<br>(44.1-71.2) | 42.4<br>[0.02 (0-0.29)] | 6.5<br>(5.6-8.8) | 88.7<br>[1.49 (0.5-3.1)] | 5.7<br>(4.4-7.4) |
| **NCM** | 11.5<br>[0.54 (0-0.28)] | 22.1<br>(18.4-26.5) | 15.6<br>[0.1 (0-0.42)] | 10.9<br>(9.6-13.5) | 19.8<br>[0.09 (0-0.44)] | 1.8<br>(1.5-2.3) | 98.8<br>[40.6 (24.6-59)] | 3.3<br>(2.2-5.3) |
| **Granulocytes** | 76.8<br>[0.015 (0.005-0.03)] | 60.5<br>(48.1-88.2) | 74.4<br>[0.019 (0.005-0.06)] | 38.7<br>(30.0-52.8) | 73.3<br>[0.025 (0.003-0.05)] | 6.8<br>(5.2-8.6) | 98.8<br>[0.414 (0.20-0.71)] | 3.7<br>(3.6-4.4) |

**Table S2.**
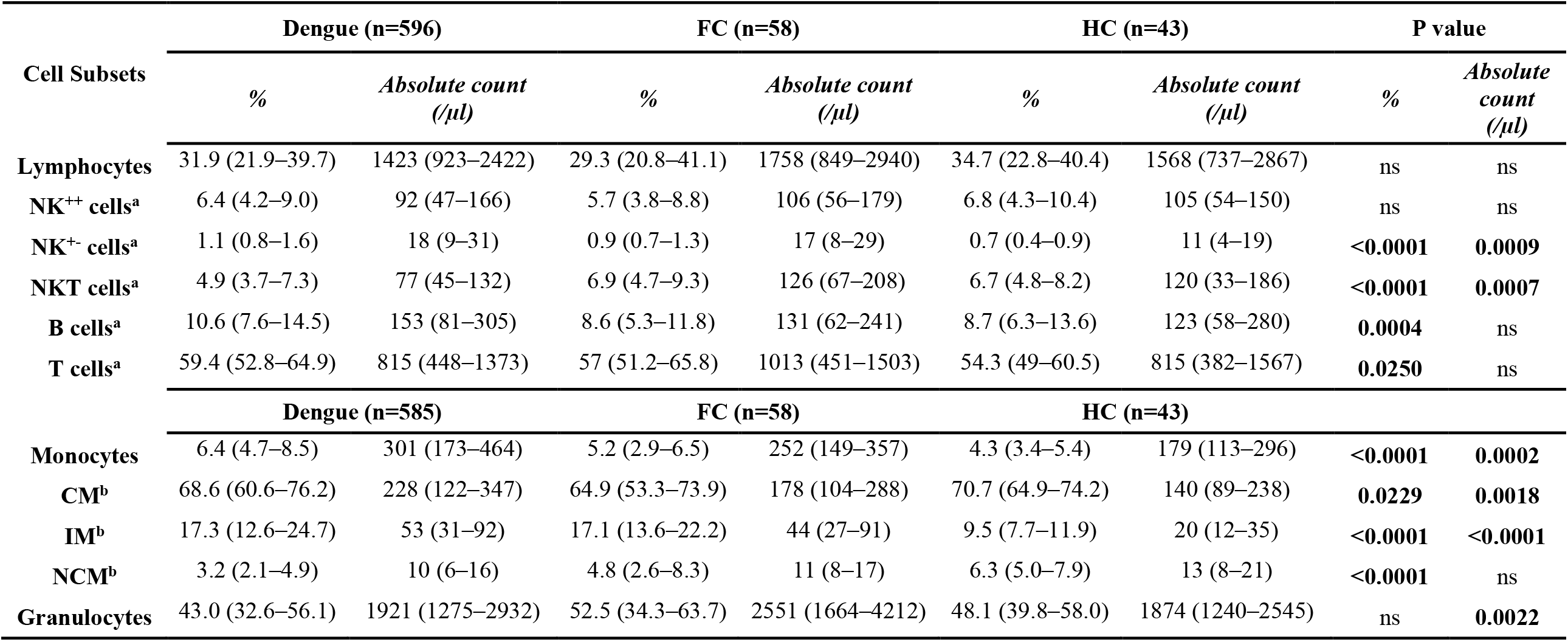
Immune cell subsets in dengue and control groups. The presented values are median (IQR). Kruskal-Wallis test followed by Bonferroni correction for multiple comparisons was performed to compare the difference of absolute count /µl blood and % of cell subsets between volunteer groups and the P values are indicated. Number of subjects in each group is given in parentheses. % of cell subsets - (^a^) represents percentage of lymphocytes and (^b^) represents percentage of monocytes. Abbreviations: FC – febrile controls; HC – healthy controls; ns - not significant; IQR – Inter Quartile Range; NKT – CD56^+^CD3^+^ cells; NK^++^ - CD56^+^CD16^+^ cells; NK^+-^ - CD56^+^CD16^-^ cells; B cells – CD19^+^ cells; T cells – CD3^+^ cells; CM – CD14^+^CD16^-^ classical monocytes; IM – CD14^+^CD16^+^ intermediate monocytes; NCM – CD14^-^CD16^+^ non-classical monocytes.

**Table S3.**
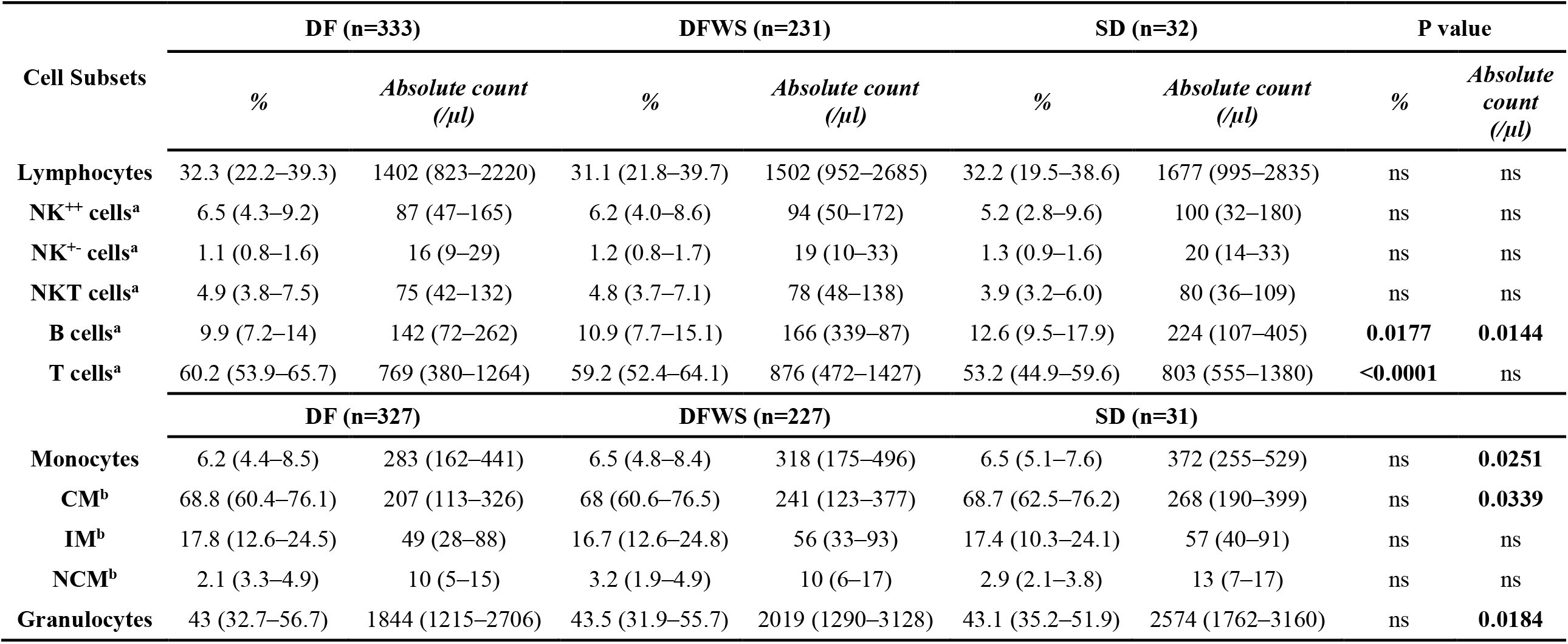
Cell subsets as a function of dengue severity. The presented values are median (IQR). Kruskal-Wallis test followed by Bonferroni correction for multiple comparisons was performed to compare the difference of absolute count/µl blood and % of cell subsets between dengue severity groups and the P values are indicated. Number of subjects in each group is given in parentheses. % of cell subsets - (^a^) represents percentage of lymphocytes and (^b^) represents percentage of monocytes. Abbreviations: DF – dengue fever; DFWS – dengue fever with warning signs; SD – severe dengue; ns - not significant; IQR – Inter Quartile Range; NKT – CD56^+^CD3^+^ cells; NK^++^ - CD56^+^CD16^+^ cells; NK^+-^ - CD56^+^CD16^-^ cells; B cells – CD19^+^ cells; T cells – CD3^+^ cells; CM – CD14^+^CD16^-^ classical monocytes; IM – CD14^+^CD16^+^ intermediate monocytes; NCM – CD14^-^ CD16^+^ non-classical monocytes.

**Table S4.** Correlation analysis of cytokines between subsets. Spearman’s correlation coefficient and P values for cytokines from various subsets are reported. Abbreviations: n – number of samples; NKT – CD56^+^CD3^+^ cells; NK^++^ - CD56^+^CD16^+^ cells; B cells – CD19^+^ cells; CM – CD14^+^CD16^-^ classical monocytes; IM – CD14^+^CD16^+^ intermediate monocytes.

|  | n | Spearman r (95% CI) | P value |
| --- | --- | --- | --- |
| TNF-$\alpha^+$ NK$^{++}$ & TNF-$\alpha^+$ NKT cells | 596 | 0.77 (0.73-0.80) | <0.0001 |
| TNF-$\alpha^+$ NK$^{++}$ & TNF-$\alpha^+$ B cells | 596 | 0.64 (0.58-0.68) | <0.0001 |
| TNF-$\alpha^+$ NK$^{++}$ & TNF-$\alpha^+$ granulocytes | 582 | 0.64 (0.58-0.68) | <0.0001 |
| TNF-$\alpha^+$ CM & TNF-$\alpha^+$ granulocytes | 585 | 0.71 (0.67-0.75) | <0.0001 |
| TNF-$\alpha^+$ IM & TNF-$\alpha^+$ granulocytes | 585 | 0.63 (0.59-0.68) | <0.0001 |
| TNF-$\alpha^+$ NK$^{++}$ & IFN-$\gamma^+$ TNF-$\alpha^+$ NKT cells | 596 | 0.73 (0.69-0.77) | <0.0001 |
| IFN-$\gamma^+$ TNF-$\alpha^+$ NKT & IFN-$\gamma^+$ TNF-$\alpha^+$ NK$^{++}$ cells | 596 | 0.76 (0.72-0.79) | <0.0001 |
| IFN-$\gamma^+$ TNF-$\alpha^+$ NKT & IFN-$\gamma^+$ NK$^{++}$ cells | 596 | 0.61 (0.56-0.66) | <0.0001 |
| IFN-$\gamma^+$ TNF-$\alpha^+$ NK$^{++}$ & IFN-$\gamma^+$ NKT cells | 596 | 0.72 (0.67-0.75) | <0.0001 |
| IFN-$\gamma^+$ NKT & IFN-$\gamma^+$ NK$^{++}$ cells | 596 | 0.71 (0.66-0.74) | <0.0001 |
| IFN-$\gamma^+$ NKT & TNF-$\alpha^+$ NK$^{++}$ cells | 596 | 0.69 (0.64-0.73) | <0.0001 |

**Table S5.** Biomarker performance of IFN-γ^+^CD56^+^CD3^+^ NKT cells, IFN-γ^+^TNF-α^+^CD56^+^CD3^+^ NKT cells and monofunctional IL-6^+^granulocytes. The tabulated values are AUC, cut-off used, sensitivity (%), specificity (%) and likelihood ratio from ROC analysis in total and homogeneous patient groups. ROC analysis was carried out between recovered DF patients and those who worsened during the study. Abbreviations: AUC – area under the curve; CI – confidence interval; ROC – receiver operating characteristic; DF – dengue fever. Cut off represents (^a^) % cytokine secreting cells or (^b^) probability obtained from regression analysis.

| Subset and cytokine | Patient groups | AUC (95% CI) | Cut-off | Sensitivity (%) | Specificity (%) | Likelihood Ratio |
| --- | --- | --- | --- | --- | --- | --- |
| **IFN-γ<sup>+</sup>CD56<sup>+</sup>CD3<sup>+</sup> NKT cells** | Total | 0.76<br>(0.64-0.88) | <1.202<sup>a</sup> | 90 | 61.56 | 2.34 |
|  | Abnormal liver enzymes | 0.83<br>(0.74-0.92) | <1.216<sup>a</sup> | 100 | 60.32 | 2.52 |
| **IFN-γ<sup>+</sup>TNF-α<sup>+</sup>CD56<sup>+</sup>CD3<sup>+</sup> NKT cells** | Total | 0.77<br>(0.64-0.90) | <1.072<sup>a</sup> | 90 | 60.96 | 2.31 |
|  | Abnormal liver enzymes | 0.83<br>(0.74-0.93) | <1.072<sup>a</sup> | 100 | 59.52 | 2.47 |
| **IL-6<sup>+</sup> Granulocytes** | Total | 0.75<br>(0.67-0.83) | <0.005<sup>a</sup> | 90 | 66.36 | 2.67 |
| **IL-6<sup>+</sup> Granulocytes, IFN-γ<sup>+</sup>CD56<sup>+</sup>CD3<sup>+</sup> NKT cells** | Total | 0.85<br>(0.78-0.92) | >0.042<sup>b</sup> | 90 | 75.93 | 3.73 |
|  | Abnormal AST | 0.90<br>(0.85-0.95) | >0.056<sup>b</sup> | 100 | 81.9 | 5.53 |
|  | Abnormal liver enzymes | 0.88<br>(0.83-0.94) | >0.51<sup>b</sup> | 100 | 75.7 | 4.12 |

**Table S6.** Antibodies used for flow cytometry.

| Target (Clone) | Species | Fluorochrome/Conjugate | Company | Catalog no. |
| --- | --- | --- | --- | --- |
| **NK cell panel – Surface Markers** |  |  |  |  |
| CD19 (HIB19) | Mouse IgG1, $\kappa$ | PE-Cy5 | BioLegend | 302210 |
| CD56 (B159) | Mouse IgG1, $\kappa$ | PE-Cy7 | BD Bioscience | 557747 |
| CD3 (SK7) | Mouse BALB/cIgG1, $\kappa$ | APC-H7 | BD Bioscience | 560176 |
| CD16 (3G8) | Mouse CDF1 IgG1, $\kappa$ | BV510 | BD Bioscience | 563830 |
| **Monocyte Panel – Surface Markers** |  |  |  |  |
| CD14 (M5E2) | Mouse IgG2a, $\kappa$ | PerCP- Cy5.5 | BD Bioscience | 550787 |
| TLR2 (11G7) | Mouse IgG1, $\kappa$ | BV510 | BD Bioscience | 742767 |
| CD16 (3G8) | Mouse CDF1 IgG1, $\kappa$ | BV605 | BD Bioscience | 563172 |
| **Intracellular Cytokine Markers** |  |  |  |  |
| IP-10 (6D4/D6/G2) | Mouse IgG2a, $\kappa$ | PE | BD Bioscience | 555049 |
| IL-10 (JES3-19F1) | Rat IgG2a, $\kappa$ | APC | BD Bioscience | 554707 |
| IFN-γ (B27) | Mouse IgG1, $\kappa$ | BV605 | BD Bioscience | 562974 |
| TNF-α (MAb11) | Mouse IgG1, $\kappa$ | BV750 | BD Bioscience | 566359 |
| IL-6 (MQ2-6A32) | Rat IgG2a, $\kappa$ | FITC | BD Bioscience | 557696 |

**Table S7.** Criteria used to define positive responses in test samples. The minimum number of events and the number of events over the control used to define a positive response in test samples for all cytokines from innate cell subsets are reported. Abbreviations: NKT – CD56^+^CD3^+^ cells; NK^++^ - CD56^+^CD16^+^ cells; NK^+-^ - CD56^+^CD16^-^ cells; B cells – CD19^+^ cells; CM – CD14^+^ classical monocytes; IM – CD14^+^CD16^+^ intermediate monocytes; NCM – CD16^+^ non-classical monocytes.

| **Cell subset** | **IFN-γ** |  | **IL-10** |  | **IP-10** |  | **TNF-α** |  |
| --- | --- | --- | --- | --- | --- | --- | --- | --- |
| *Events* | *Minimum* | *over control* | *Minimum* | *over control* | *Minimum* | *over control* | *Minimum* | *over control* |
| NKT | 15 | twice | 10 | 10 | 10 | 10 | 15 | twice |
| NK<sup>++</sup> | 15 | 15 | 10 | 10 | 10 | 10 | 15 | twice |
| NK<sup>+-</sup> | 15 | twice | 10 | 10 | 10 | 10 | 15 | 10 |
| B | 15 | 10 | 10 | 10 | 10 | 10 | 15 | 15 |
|  | **IL-6** |  | **IL-10** |  | **IP-10** |  | **TNF-α** |  |
| CM | 10 | 10 | 10 | 10 | 10 | 10 | 15 | 15 |
| IM | 10 | 10 | 10 | 10 | 10 | 10 | 15 | 15 |
| NCM | 10 | 10 | 10 | 10 | 10 | 10 | 15 | twice |
| Granulocytes | 15 | 15 | 15 | 15 | 15 | 15 | 15 | twice |

**Table S8.** Total number of cytokine-secreting innate immune cell events from control and test samples. The median, IQR and range are reported. Abbreviations: IQR – Inter Quartile Range; CM – CD14^+^ classical monocytes; IM – CD14^+^CD16^+^ intermediate monocytes; NCM – CD16^+^ non-classical monocytes NKT – CD56^+^CD3^+^ cells; NK^++^ - CD56^+^CD16^+^ cells; NK^+-^ - CD56^+^CD16^-^ cells; B cells – CD19^+^ cells.

| Cell Subsets | Events | TNF-α | | IL-6 | | IP-10 | | IL-10 | |
| --- | --- | --- | --- | --- | --- | --- | --- | --- | --- |
|  |  | Control | Test | Control | Test | Control | Test | Control | Test |
| CM | Median | 4 | 70 | 5 | 9 | 5 | 19 | 10 | 17 |
|  | IQR | 2-8 | 33-128 | 2-17 | 4-28 | 1-20 | 4-78 | 3-35 | 7-44 |
|  | Range | 0-122 | 0-1557 | 0-402 | 0-1066 | 0-727 | 0-2388 | 0-759 | 0-1146 |
| IM | Median | 37 | 191 | 15 | 21 | 18 | 24 | 31 | 37 |
|  | IQR | 16-89 | 86-341 | 8-29 | 9-45 | 8-33 | 12-47 | 17-65 | 16-79 |
|  | Range | 1-2044 | 3-3312 | 1-843 | 0-709 | 0-1191 | 0-507 | 1-441 | 0-711 |
| NCM | Median | 2 | 564 | 2 | 3 | 1 | 4 | 4 | 6 |
|  | IQR | 1-4 | 324-927 | 0-5 | 1-8 | 0-4 | 1-9 | 1-10 | 2-15 |
|  | Range | 0-385 | 0-2686 | 0-107 | 0-205 | 0-66 | 0-229 | 0-107 | 0-157 |
| Granulocytes | Median | 38 | 1272 | 21 | 76 | 23 | 91 | 92 | 180 |
|  | IQR | 23-66 | 637-2136 | 9-47 | 40-138 | 11-49 | 40-223 | 56-147 | 110-268 |
|  | Range | 4-1565 | 38-31503 | 0-709 | 1-969 | 0-883 | 2-3534 | 9-1210 | 10-1079 |
| | | TNF-α | | IFN-γ | | IP-10 | | IL-10 | |
|  |  | Control | Test | Control | Test | Control | Test | Control | Test |
| NKT | Median | 6 | 364 | 4 | 160 | 2 | 12 | 4 | 11 |
|  | IQR | 3-13 | 186-665 | 1-8 | 81-310 | 0-7 | 4-35 | 1-9 | 4-23 |
|  | Range | 0-188 | 0-2578 | 0-129 | 1-2786 | 0-376 | 0-995 | 0-65 | 0-1032 |
| NK++ | Median | 1 | 131 | 3 | 44 | 2 | 13 | 2 | 4 |
|  | IQR | 0-2 | 63-253 | 1-8 | 22-86 | 0-5 | 4-47 | 0-6 | 1-10 |
|  | Range | 0-22 | 1-1977 | 0-85 | 0-1147 | 0-305 | 0-3825 | 0-46 | 0-59 |
| NK+- | Median | 0 | 12 | 0 | 4 | 0 | 2 | 0 | 1 |
|  | IQR | 0-1 | 5-25 | 0-0 | 1-10 | 0-1 | 0-9 | 0-1 | 0-2 |
|  | Range | 0-31 | 0-226 | 0-135 | 0-1188 | 0-335 | 0-383 | 0-18 | 0-49 |
| B | Median | 17 | 118 | 7 | 26 | 2 | 10 | 4 | 9 |
|  | IQR | 8-37 | 53-251 | 2-18 | 11-54 | 1-7 | 3-32 | 2-9 | 5-17 |
|  | Range | 0-398 | 0-1764 | 0-108 | 0-430 | 0-318 | 0-304 | 0-61 | 0-92 |

**Table S9.** Gender distribution among dengue patients. Numbers of total dengue patients admitted and those recruited for our study (between June 24, 2019 and November 29, 2019) in each hospital are listed according to gender. Abbreviations: N – Number of patients; % - percentage of male/female patients; KIMS - Kempegowda Institute of Medical Sciences; BMCRI - Bangalore Medical College and Research Institute; SJMC - St. John’s Medical College; RMCH - M S Ramaiah Medical College and Hospital.

| **Dengue Patients Recruited** |  |  |  |  |  |  |  |  |  |  |
| --- | --- | --- | --- | --- | --- | --- | --- | --- | --- | --- |
|  | **KIMS** |  | **BMCRI** |  | **RMCH** |  | **SJMC** |  | **Total** |  |
|  | **N** | **%** | **N** | **%** | **N** | **%** | **N** | **%** | **N** | **%** |
| **Male** | 147 | 80 | 65 | 65 | 80 | 60 | 136 | 76 | 428 | 72 |
| **Female** | 36 | 20 | 35 | 35 | 53 | 40 | 44 | 24 | 168 | 28 |
| **Total** | 183 |  | 100 |  | 133 |  | 180 |  | 596 |  |
| **Total Dengue Admissions** |  |  |  |  |  |  |  |  |  |  |
|  | **KIMS** |  | **BMCRI** |  | **RMCH** |  | **SJMC** |  | **Total** |  |
|  | **N** | **%** | **N** | **%** | **N** | **%** | **N** | **%** | **N** | **%** |
| **Male** | 2383 | 68 | 935 | 54 | 642 | 60 | 1280 | 59 | 5238 | 62 |
| **Female** | 1139 | 32 | 798 | 46 | 429 | 40 | 898 | 41 | 3265 | 38 |
| **Total** | 3522 |  | 1733 |  | 1071 |  | 2178 |  | 8503 |  |

**Table S10.** Demographic characteristics and clinical symptoms of dengue patients. Age and days of fever at presentation are represented as median (IQR). Statistical differences in proportion of gender, clinical symptoms and primary/secondary infection between dengue patient groups based on severity was determined by either Chi-Square or Fisher’s exact test. P values are tabulated; ^a^ - Kruskal-Wallis test; * - Fisher’s exact test. Abbreviations: dpso – days post symptom onset; IQR – Inter Quartile Range; n – number of patients; DF – dengue fever; DFWS – dengue fever with warning signs; SD – severe dengue.

| Variable | DF<br>(n=333) | DFWS<br>(n=231) | SD<br>(n=32) | P value |  |  |
| --- | --- | --- | --- | --- | --- | --- |
|  |  |  |  | DF-DFWS | DF- SD | DFWS-SD |
| Age; median<br>(IQR) | 26.8<br>(21.8, 35.04) | 28<br>(22, 36.3) | 29<br>(26.1, 43) |  | 0.098<sup>a</sup> |  |
| dpso;<br>median(IQR) | 4<br>(3,5) | 4<br>(4,5) | 4<br>(3,5) |  | **0.0008<sup>a</sup>** |  |
| Male : Female | 238:95 | 164:76 | 26:6 | 0.902 | 0.238 | 0.225 |
| Nausea; n (%) | 127 (38.1) | 127 (55) | 18 (56.3) | **<0.0005** | **0.046** | 0.89 |
| Head ache; n (%) | 155 (46.5) | 132 (57.1) | 16 (50) | **0.013** | 0.708 | 0.446 |
| Abdomen Pain; n (%) | 0 (0) | 109 (18.3) | 16 (50) | **<0.0005** | **<0.0005\*** | 0.76 |
| Body Pain; n (%) | 171 (51.4) | 146 (63.2) | 14 (43.8) | **0.005** | 0.411 | 0.035 |
| Rashes; n (%) | 22(6.6) | 25(10.8) | 2(6.3) | **0.075** | 1* | 0.549* |
| Splenomegaly; n (%) | 3(0.9) | 8(3.5) | 2(6.3) | **0.03\*** | **0.063\*** | 0.349* |
| Hepatomegaly; n (%) | 0 | 24(10.5) | 4(12.5) | **<0.0005** | **<0.0005<sup>a</sup>** | 0.536* |
| Petechiae; n (%) | 1(0.3) | 13(5.6) | 0 | **<0.0005** | 1* | 0.378* |
| Ascites; n (%) | 0 | 39(16.9) | 7(21.9) | **<0.0005** | **<0.0005\*** | 0.486 |
| Primary : Secondary | 214:118 | 109:120 | 13:18 | **<0.0005** | **0.008** | 0.457 |
| Hemorrhage; n (%) | 2 (0.6) | 47 (20) | 21 (65) | **<0.0001\*** | **<0.0001\*** | **<0.0001\*** |

**Table S11.** Clinical laboratory parameters of dengue patients. Hemoglobin and highest hematocrit are represented as mean (standard deviation); Lowest platelet count, albumin, aspartate transaminase and alanine transaminase as median (IQR). Based on normality test, ANOVA test was used for parametric data and Kruskal-Wallis test for non-parametric data to find significant difference across dengue severity based on clinical parameters. Abbreviations: IQR – Inter Quartile Range; DF – dengue fever; DFWS – dengue fever with warning signs; SD –severe dengue.

| **Variable** | **DF**<br>**(n=333)** | **DFWS**<br>**(n=231)** | **SD**<br>**(n=32)** | **P value** | **Statistical test** |
| --- | --- | --- | --- | --- | --- |
| Hemoglobin at enrollment; mean (SD) | 14.43<br>(1.95) | 14.48<br>(2.1) | 15.14<br>(2.25) | 0.10 | ANOVA |
| Lowest Platelet count (x10<sup>3</sup>); median (IQR) | 50<br>(27,85) | 29<br>(14-56) | 16<br>(10,32) | **<0.0001** | Kruskal-Wallis test |
| Highest Hematocrit %; mean (SD) | 42.9<br>(5.23) | 43.84<br>(5.5) | 44.49<br>(6.4) | 0.09 | ANOVA |
| Albumin g/dL; median (IQR) | 3.8<br>(3.5,4.1) | 3.6<br>(3.3,4) | 3.4<br>(3.2,3.7) | **<0.0001** | Kruskal-Wallis test |
| Aspartate transaminase IU/L; median (IQR) | 86<br>(50,143) | 114<br>(75,189) | 123.5<br>(91,880) | **<0.0001** | Kruskal-Wallis test |
| Alanine transaminase IU/L; median (IQR) | 56<br>(29.5,95.5) | 70<br>(45, 118) | 98<br>(51, 265) | **<0.0001** | Kruskal-Wallis test |

| Section & Topic | No | Item | Reported on page # |
| --- | --- | --- | --- |
| **TITLE OR ABSTRACT** |  |  |  |
|  | **1** | Identification as a study of diagnostic accuracy using at least one measure of accuracy (such as sensitivity, specificity, predictive values, or AUC) | Main page 1,2 |
| **ABSTRACT** |  |  |  |
|  | **2** | Structured summary of study design, methods, results, and conclusions (for specific guidance, see STARD for Abstracts) | Main page 1,2 |
| **INTRODUCTION** |  |  |  |
|  | **3** | Scientific and clinical background, including the intended use and clinical role of the index test | Main page 2,3 |
|  | **4** | Study objectives and hypotheses | Main page 2,3 |
| **METHODS** |  |  |  |
| *Study design* | **5** | Whether data collection was planned before the index test and reference standard were performed (prospective study) or after (retrospective study) | Supplementary page 2,3 |
| *Participants* | **6** | Eligibility criteria | Supplementary page 2,3 |
|  | **7** | On what basis potentially eligible participants were identified (such as symptoms, results from previous tests, inclusion in registry) | Supplementary page 2,3 |
|  | **8** | Where and when potentially eligible participants were identified (setting, location and dates) | Supplementary page 2 |
|  | **9** | Whether participants formed a consecutive, random or convenience series | Supplementary page 2 |
| *Test methods* | **10a** | Index test, in sufficient detail to allow replication | Supplementary page 4,5 |
|  | **10b** | Reference standard, in sufficient detail to allow replication | Supplementary page 2 |
|  | **11** | Rationale for choosing the reference standard (if alternatives exist) | n/a |
|  | **12a** | Definition of and rationale for test positivity cut-offs or result categories of the index test, distinguishing pre-specified from exploratory | n/a |
|  | **12b** | Definition of and rationale for test positivity cut-offs or result categories of the reference standard, distinguishing pre-specified from exploratory | n/a |
|  | **13a** | Whether clinical information and reference standard results were available to the performers/readers of the index test | Supplementary pages 2, 5 |
|  | **13b** | Whether clinical information and index test results were available to the assessors of the reference standard | Supplementary page 5 |
| *Analysis* | **14** | Methods for estimating or comparing measures of diagnostic accuracy | Supplementary page 5,6 |
|  | **15** | How indeterminate index test or reference standard results were handled | Supplementary page 3 |
|  | **16** | How missing data on the index test and reference standard were handled | Supplementary page 3 |
|  | **17** | Any analyses of variability in diagnostic accuracy, distinguishing pre-specified from exploratory | n/a |
|  | **18** | Intended sample size and how it was determined | Supplementary page 2 |
| **RESULTS** |  |  |  |
| *Participants* | **19** | Flow of participants, using a diagram | Supplementary page 9 |
|  | **20** | Baseline demographic and clinical characteristics of participants | Supplementary page 37,38 |
|  | **21a** | Distribution of severity of disease in those with the target condition | Supplementary page 37,38 |
|  | **21b** | Distribution of alternative diagnoses in those without the target condition | n/a |
|  | **22** | Time interval and any clinical interventions between index test and reference standard | n/a |
| *Test results* | **23** | Cross tabulation of the index test results (or their distribution) by the results of the reference standard | Main page 5,6 |
|  | **24** | Estimates of diagnostic accuracy and their precision (such as 95% confidence intervals) | Supplementary page 31 |
|  | **25** | Any adverse events from performing the index test or the reference standard | n/a |
| **DISCUSSION** |  |  |  |
|  | **26** | Study limitations, including sources of potential bias, statistical uncertainty, and generalisability | Main page 11,12 |
|  | **27** | Implications for practice, including the intended use and clinical role of the index test | Main page 11,12 |
| **OTHER INFORMATION** |  |  |  |
|  | **28** | Registration number and name of registry | n/a |
|  | **29** | Where the full study protocol can be accessed | n/a |
|  | **30** | Sources of funding and other support; role of funders | Main page 15 |

## AIM

STARD stands for “Standards for Reporting Diagnostic accuracy studies”. This list of items was developed to contribute to the completeness and transparency of reporting of diagnostic accuracy studies. Authors can use the list to write informative study reports. Editors and peer-reviewers can use it to evaluate whether the information has been included in manuscripts submitted for publication.

## EXPLANATION

A **diagnostic accuracy study** evaluates the ability of one or more medical tests to correctly classify study participants as having a **target condition**. This can be a disease, a disease stage, response or benefit from therapy, or an event or condition in the future. A medical test can be an imaging procedure, a laboratory test, elements from history and physical examination, a combination of these, or any other method for collecting information about the current health status of a patient.

The test whose accuracy is evaluated is called **index test**. A study can evaluate the accuracy of one or more index tests. Evaluating the ability of a medical test to correctly classify patients is typically done by comparing the distribution of the index test results with those of the **reference standard**. The reference standard is the best available method for establishing the presence or absence of the target condition. An accuracy study can rely on one or more reference standards.

If test results are categorized as either positive or negative, the cross tabulation of the index test results against those of the reference standard can be used to estimate the **sensitivity** of the index test (the proportion of participants *with* the target condition who have a positive index test), and its **specificity** (the proportion *without* the target condition who have a negative index test). From this cross tabulation (sometimes referred to as the contingency or “2×2” table), several other accuracy statistics can be estimated, such as the positive and negative **predictive values** of the test. Confidence intervals around estimates of accuracy can then be calculated to quantify the statistical **precision** of the measurements.

If the index test results can take more than two values, categorization of test results as positive or negative requires a **test positivity cut-off**. When multiple such cut-offs can be defined, authors can report a receiver operating characteristic (ROC) curve which graphically represents the combination of sensitivity and specificity for each possible test positivity cut-off. The **area under the ROC curve** informs in a single numerical value about the overall diagnostic accuracy of the index test.

The **intended use** of a medical test can be diagnosis, screening, staging, monitoring, surveillance, prediction or prognosis. The **clinical role** of a test explains its position relative to existing tests in the clinical pathway. A replacement test, for example, replaces an existing test. A triage test is used before an existing test; an add-on test is used after an existing test.

Besides diagnostic accuracy, several other outcomes and statistics may be relevant in the evaluation of medical tests. Medical tests can also be used to classify patients for purposes other than diagnosis, such as staging or prognosis. The STARD list was not explicitly developed for these other outcomes, statistics, and study types, although most STARD items would still apply.

## DEVELOPMENT

This STARD list was released in 2015. The 30 items were identified by an international expert group of methodologists, researchers, and editors. The guiding principle in the development of STARD was to select items that, when reported, would help readers to judge the potential for bias in the study, to appraise the applicability of the study findings and the validity of conclusions and recommendations. The list represents an update of the first version, which was published in 2003.

More information can be found on http://www.equator-network.org/reporting-guidelines/stard.

